# Immune Responses to COVID-19 mRNA Vaccines in Patients with Solid Tumors on Active, Immunosuppressive Cancer Therapy

**DOI:** 10.1101/2021.05.13.21257129

**Authors:** Rachna T. Shroff, Pavani Chalasani, Ran Wei, Daniel Pennington, Grace Quirk, Marta V. Schoenle, Kameron L. Peyton, Jennifer L. Uhrlaub, Tyler J. Ripperger, Mladen Jergović, Shelby Dalgai, Alexander Wolf, Rebecca Whitmer, Hytham Hammad, Amy Carrier, Aaron J. Scott, Janko Nikolich-Žugich, Michael Worobey, Ryan Sprissler, Michael Dake, Bonnie J. LaFleur, Deepta Bhattacharya

## Abstract

Vaccines against SARS-CoV-2 have shown high efficacy, but immunocompromised participants were excluded from controlled clinical trials. We compared immune responses to the Pfizer/BioNTech mRNA vaccine in solid tumor patients (n=53) on active cytotoxic anti-cancer therapy to a control cohort (n=50) as an observational study. Using live SARS-CoV-2 assays, neutralizing antibodies were detected in 67% and 80% of cancer patients after the first and second immunizations, respectively, with a 3-fold increase in median titers after the booster. Similar trends were observed in serum antibodies against the receptor-binding domain (RBD) and S2 regions of Spike protein, and in IFN*γ*+ Spike-specific T cells. Yet the magnitude of each of these responses was diminished relative to the control cohort. We therefore quantified RBD- and Spike S1-specific memory B cell subsets as predictors of anamnestic responses to additional immunizations. After the second vaccination, Spike-specific plasma cell-biased memory B cells were observed in most cancer patients at levels similar to those of the control cohort after the first immunization. We initiated an interventional phase 1 trial of a third booster shot (NCT04936997); primary outcomes were immune responses with a secondary outcome of safety. After a third immunization, the 20 participants demonstrated an increase in antibody responses, with a median 3-fold increase in virus-neutralizing titers. Yet no improvement was observed in T cell responses at 1 week after the booster immunization. There were mild adverse events, primarily injection site myalgia, with no serious adverse events after a month of follow-up. These results suggest that a third vaccination improves humoral immunity against COVID-19 in cancer patients on active chemotherapy with no severe adverse events.

---

The COVID-19 pandemic has led to over 200 million infections worldwide and claimed over 4 million lives to date. While non-pharmaceutical public health interventions managed to control outbreaks in certain countries, most of the global population will depend upon vaccines to mitigate the pandemic. Since the identification of SARS-CoV-2 as the causative agent of COVID-19 in January 2020^1, 2^, vaccines with very high efficacy have been developed and deployed with remarkable speed. Independent clinical trials demonstrated 94-95% vaccine efficacy against symptomatic disease caused by SARS-CoV-2 for both the Pfizer/BioNTech and Moderna mRNA-based vaccines^3, 4^. Based on these data, in December 2020, both the Pfizer/BioNTech and Moderna vaccines were granted emergency use authorization by regulatory agencies in the United Kingdom and North America. Subsequent observational studies after authorization have shown that these vaccines also have high effectiveness against asymptomatic infections and suppress viral loads in breakthrough infections^5–8^. These data portend a marked overall reduction in community transmission once widespread vaccination is achieved.

These clinical trials, however, largely excluded immunocompromised individuals, including patients on immunosuppressive therapies to control chronic inflammatory conditions, primary immunodeficiencies, organ transplant recipients, and cancer patients on cytotoxic chemotherapy. As the number of deaths from this devastating virus has exceeded 4 million, concern has been high about its impact on cancer patients. This is especially true since a study from the COVID-19 Cancer Consortium showed a 13% 30-day all-cause mortality from COVID-19 in a study of 928 patients^9^. Importantly, the investigators noted a higher risk of death in patients with active cancer^9^.

Beyond the obvious direct benefits to these patients, vaccine-induced protection of immunocompromised individuals is of substantial indirect benefit to the general population. Some highly transmissible SARS-CoV-2 variants of concern that partially evade antibody responses are suspected to have arisen following prolonged evolution within immunocompromised patients^10–15^. Even partial vaccine-induced immunity is likely to reduce within-host viral population size and duration of within-host viral persistence and evolution, thereby slowing the emergence of future problematic variants^16^. Yet protective immune correlates of antibodies and memory B and T cells remain to be quantitatively defined^17^. Thus, optimal strategies are needed to elevate post-vaccination immunity in vulnerable immunocompromised populations to similar levels observed in healthy individuals. For individuals who cannot mount such an immune response, widespread community vaccination and targeted strategies to immunize close contacts will be required for indirect protection.

Several recent reports have shown diminished immune responses to SARS-CoV-2 infections and mRNA vaccines in subsets of immunocompromised patients, though these vary greatly with the nature of the immunosuppressive therapy. For example, patients with autoimmune conditions or chronic lymphocytic leukemia treated with B cell-depleting antibodies have predictably diminished humoral responses to vaccination^18^, whereas responses by patients on anti-TNF⍺ therapies are less affected^19^. As another example, organ transplant recipients mount very poor antibody responses to the first mRNA immunization relative to healthy individuals^20^, but improve somewhat after the second immunization^21^. Similarly, in cancer patients with solid or hematological malignancies, antibody responses are also markedly diminished after the first immunization but often improve after the second^22^. Yet because a relatively small group of these cancer patients was tested after the second immunization and because this group contained a mixture of those on cytotoxic chemotherapy and checkpoint blockade therapy, more data are required to instruct how best to protect this vulnerable population.

We followed serological and cellular immune responses following mRNA vaccination of solid tumor patients on active cytotoxic chemotherapy. After the first immunization, we observed a higher fraction of patients with neutralizing antibodies than had previously been reported. Both the magnitude and frequency of these and T cell responses improved after the second vaccination but did not reach the levels observed in our control cohort. After the second dose, Spike RBD and other S1-specific memory B cells were observed in cancer patients at levels similar to those observed in healthy individuals after the first immunization. These data suggested that a third immunization may substantially benefit those who mount weak antibody responses.

We therefore initiated an interventional trial for a third booster shot in this cohort. We observed improvements in overall Spike RBD-specific antibody levels and in virus-neutralizing titers. Yet no such improvement was observed in T cell responses. Reported adverse events were generally mild and similar to those reported after the second immunization. Together, these data suggest that booster vaccinations improve humoral immunity in cancer patients on chemotherapy.

## Results

Fifty-three patients with a known diagnosis of a solid tumor malignancy on active immunosuppressive cancer therapy were enrolled through the University of Arizona Cancer Center during their routine care. Fifty-four participants in the control cohort were enrolled through the State of Arizona’s COVID-19 vaccine point of distribution site at the University of Arizona during the phase 1B vaccination program while in the observational waiting area after their first vaccine shot and the observation (Table 1). While Table 1 contains a grouping of chemotherapeutic regimes, a full listing is included in (Extended Data Table 1).

**Table 1:** Characteristics of cohorts.

|  | **Cancer Cohort**<br>**(N=53)** | **Control Cohort**<br>**(N=50)** | **Interventional Cohort**<br>**(N=20)** |
| --- | --- | --- | --- |
| **Age** |  |  |  |
| Mean (SD) | 63.7 (9.14) | 41.3 (17.1) | 63.1 (10.1) |
| **Gender** |  |  |  |
| Female | 42 (79.2%) | 33 (66.0%) | 15 (75.0%) |
| Male | 11 (20.8%) | 17 (34.0%) | 5 (25.0%) |
| **Prednisone** |  |  |  |
| Yes | 1.00 (1.9%) | 0 (0%) | 0 (0%) |
| No | 52.0 (98.1%) | 50 (100%) | 20 (100%) |
| **Recent Surgery** |  |  |  |
| Yes | 2 (3.8%) | 0 (0%) | 1 (5.0%) |
| No | 50 (96.2%) | 50 (100%) | 19 (95.0%) |
| **Other Vaccines** |  |  |  |
| Yes | 0 (0%) | 0 (0%) | 0 (0%) |
| No | 53 (100%) | 50 (100%) | 20 (100%) |
| **Prior COVID Infection** |  |  |  |
| Yes | 4 (5.8%) | 1.00 (2.0%) | 3 (15.0%) |
| No | 49 (94.2%) | 49 (98.0%) | 17 (85.0%) |
| **Radiation** |  |  |  |
| Yes | 19 (35.8%) |  | 7 (35.0%) |
| No | 34 (64.2%) |  | 13 (65.0%) |
| Missing | 1 (1.9%) |  | 0 (0%) |
| **Days Since Treatment Prior to Draw 1** |  |  |  |
| Mean (SD) | 16.3 (51.2) |  | 5.05 (8.30) |
| **Days Since Treatment Prior to Draw 2** |  |  |  |
| Mean (SD) | 8.02 (7.64) |  | 5.65 (5.71) |
| **Days Since Treatment Prior to Draw 3** |  |  |  |
| Mean (SD) | 15.8 (8.09) |  | 7.35 (5.62) |
| **Days Since Treatment Prior to Draw 4** |  |  |  |
| Mean (SD) |  |  | 14.9 (18.6) |
| **Tumor Type** |  |  |  |
| Gastroesophageal cancer | 3 (5.8%) |  |  |
| Pancreatic cancer | 11 (21.1%) |  | 8 (40.0%) |
| Biliary cancer | 4 (7.6%) |  | 3 (15.0%) |
| Colorectal cancer | 9 (17.3%) |  | 4 (20.0%) |
| Breast cancer | 22 (42.3%) |  | 5 (25.0%) |
| Sarcoma | 1 (1.9%) |  |  |
| Ovarian cancer | 1 (1.9%) |  |  |
| **Chemotherapy\*** |  |  |  |
| Anthracycline-based | 2 (4.0%) |  | 0 (0%) |
| Fluoropyrimidine-based | 14 (26.4%) |  | 7 (35%) |
| Gemcitabine-based | 13 (24.5%) |  | 10 (50%) |
| Oral CDK4/6-based | 10 (18.9%) |  | 3 (15.0%) |
| Other Targeted Cytotoxics | 4 (7.0%) |  | 2 (10.0%) |
| Taxane/other antimicrotubule-based | 9 (17.0%) |  | 0 (0%) |

\*not mutually exclusive therefore don't sum to 100%

Blood samples for serological and cellular analyses were collected at the time of the first immunization, at the time of the second immunization, and again 5-11 days after the second vaccination (Figure 1a). Overall peripheral blood mononuclear cell (PBMC) counts were similar between the cancer and control cohorts (Extended Data Figures 1, 2a). However, we noted a reduction in the frequency of CD19+ B cells and an increase in CD13+ myeloid cells in the cancer cohort relative to controls (Extended Data Figures 2b-c). Despite the overall reduction in the frequency of B cells in the cancer cohort, naive and other activated subsets were well-represented within these B cells (Extended Data Figure 2d). Using serum from each of these samples, we first obtained single-dilution semi-quantitative data on Spike protein-specific antibody levels. Of control cohort participants, using a University of Arizona clinical serology test^23^, four tested as positive for prior SARS-CoV-2 exposure before vaccination. These participants were excluded from further analyses. For both the control and cancer cohorts, we observed progressive increases after the first and second vaccinations in antibodies specific for the S2 region of Spike protein (Figure 1b). This region contains several antibody epitopes that are conserved across other common human ꞵ-coronaviruses^25–28^, including at least one weakly neutralizing epitope^29, 30^. Although both the cancer and control cohorts showed responses, median S2-specific antibody values were diminished in cancer patients relative to the control cohort at matched timepoints (Figure 1b). As most neutralizing and protective antibodies are directed to the receptor binding domain (RBD) of Spike protein^31, 32^, we also semi-quantitatively determined the relative levels of these antibodies. Increases were also seen for RBD antibodies in both the healthy and cancer cohorts after each vaccination (Figure 1c). Yet as with antibodies against S2, the levels of RBD antibodies at draws 2 and 3 in the cancer cohort were diminished relative to healthy controls (Figure 1c). To obtain more quantitative information, we performed a full dilution series to determine antibody titers against RBD (Extended Data Figure 2e). Consistent with the semi-quantitative results, RBD antibody titers increased after the second immunization in both groups, but the median titers observed in the cancer cohort were reduced by >11-fold relative to healthy controls (Figure 1d). Seven of the cancer cohort, but none of the control cohort, failed to generate RBD-specific antibody titers above the limit of detection.

**Figure 1:**
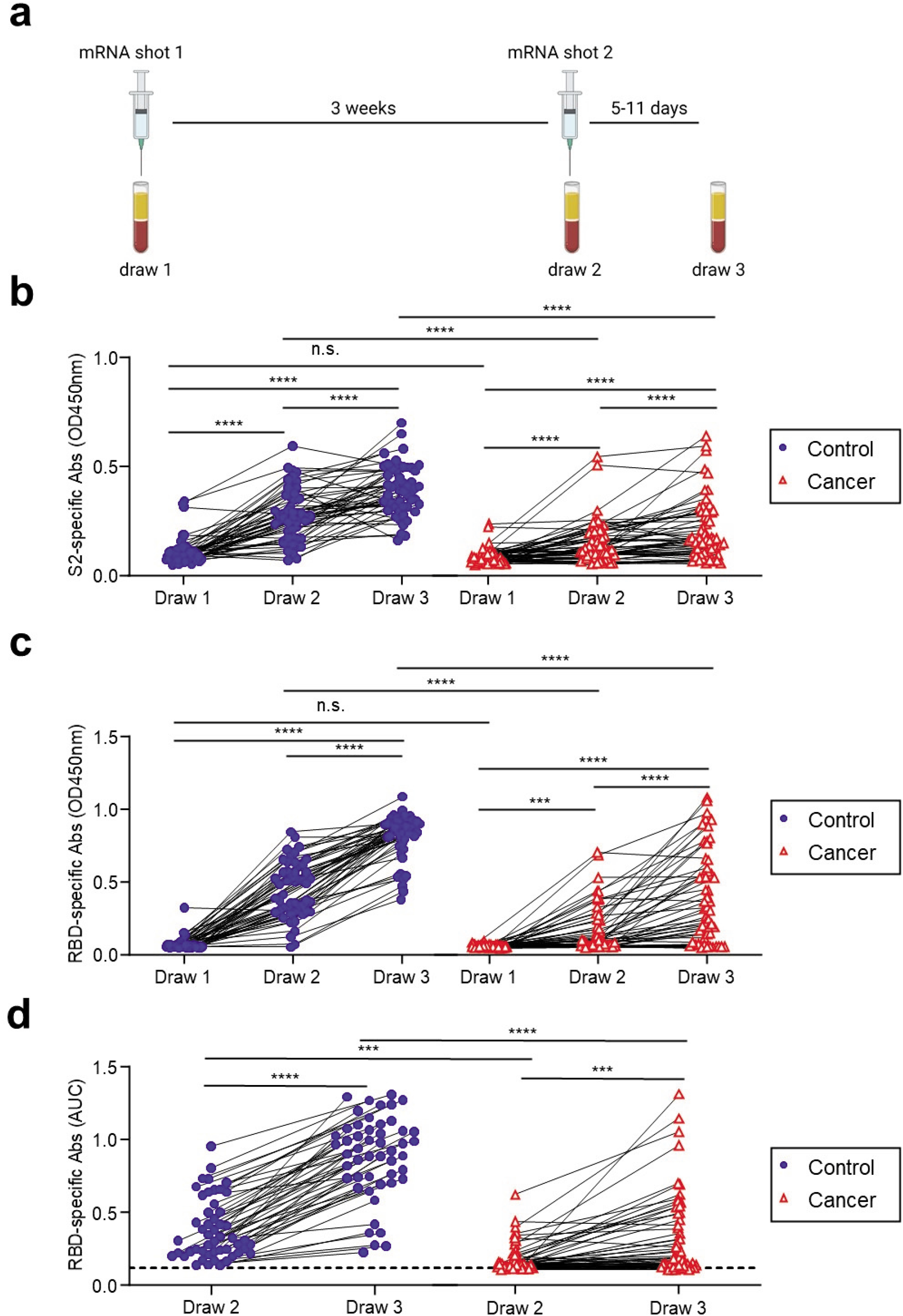
Antibody responses of cancer and control cohorts to mRNA vaccination. **a,** Schematic of blood collection (draws) after vaccination. **b,** Semi-quantitative 1:40 serum dilution ELISA results for reactivity to the S2 region of SARS-CoV-2 Spike protein. Lines connect the same individual across timepoints. Repeated measures ANOVA examines the differences in slopes between cohorts, independently from the mean differences that were demonstrated at draw 3 between cohorts. There is a statistically significant difference in slopes between cancer and control cohorts (p < 0.0001) and the average rate of change is increasing at a steeper rate in the control cohort. These paired rates between draws by cohort are statistically different in the control compared to the cancer cohort for both draw 1 and draw 2, though it is not different between draw 2 and draw 3 (p-values < 0.0001 and 0.2945, respectively). **c,** Semi-quantitative 1:40 serum dilution ELISA results for reactivity to the receptor binding domain (RBD) of SARS-CoV-2 spike protein. Lines connect the same individual at each blood draw. There is a statistically significant difference in slopes between cancer and control cohorts (p < 0.0001) and the average rate of change is steeper in the control cohort. These paired rates between draws are statistically different in the control compared to the cancer cohort for both draw 1 and draw 2 and draw 2 and draw 3 (p-values < 0.0001 and 0.0043, respectively). **d,** Quantitative titers of RBD antibodies in control and cancer cohorts. A serum concentration beginning at 1:80 was serially diluted 1:4 and area under the curve (AUC) values calculated. Lines connect the same individual across timepoints. There is a statistically significant difference between draw 2 and draw 3 between cancer and control cohorts (p < 0.0001) and the average rate of change is at a steeper increase in the control cohort. ***p<0.001; ****p<0.0001 by repeated measures ANOVA.

For most vaccines, neutralizing antibody titers are the best correlate of protection from infections^33^. We therefore directly assessed antibody-mediated neutralization of authentic live SARS-CoV-2 (WA1 isolate) after the first and second immunizations, as these assays tend to be more sensitive than those using pseudoviruses^34–37^. After the first shot, we observed a median plaque reduction neutralization test (PRNT)-90 titer of 60 in the control cohort and 20 in the cancer cohort (Figure 2). However, whereas all but one participant in the control cohort showed detectable virus neutralizing activity, this was observed in only 67% of the cancer cohort (Figure 2). After the second immunization, all healthy controls had virus-neutralizing antibodies, with a median PRNT90 titer of 540 (Figure 2). In contrast, 80% of the cancer cohort had detectable neutralizing antibodies with a median titer of 60 (Figure 2). Virus-neutralizing titers correlated with overall RBD-specific antibodies (Extended Data Figure 3). These results demonstrated that most of the cancer cohort generated protective antibodies, but at levels well below that of the control cohort after the second vaccine dose. We did not find any obvious clinical characteristics that would have modified the relationship between immunosuppression and vaccine response. Of the non-responders, 60% were breast cancer patients, 90% were female and the median age was 64. While there is no statistical power to compare this subgroup to the overall cancer cohort, the only obvious difference was treatment timing (Table 1), as the average time between treatment and vaccine dose 2 was over 2 weeks in the overall group compared to less than 1 week in non-responders.

**Figure 2:**
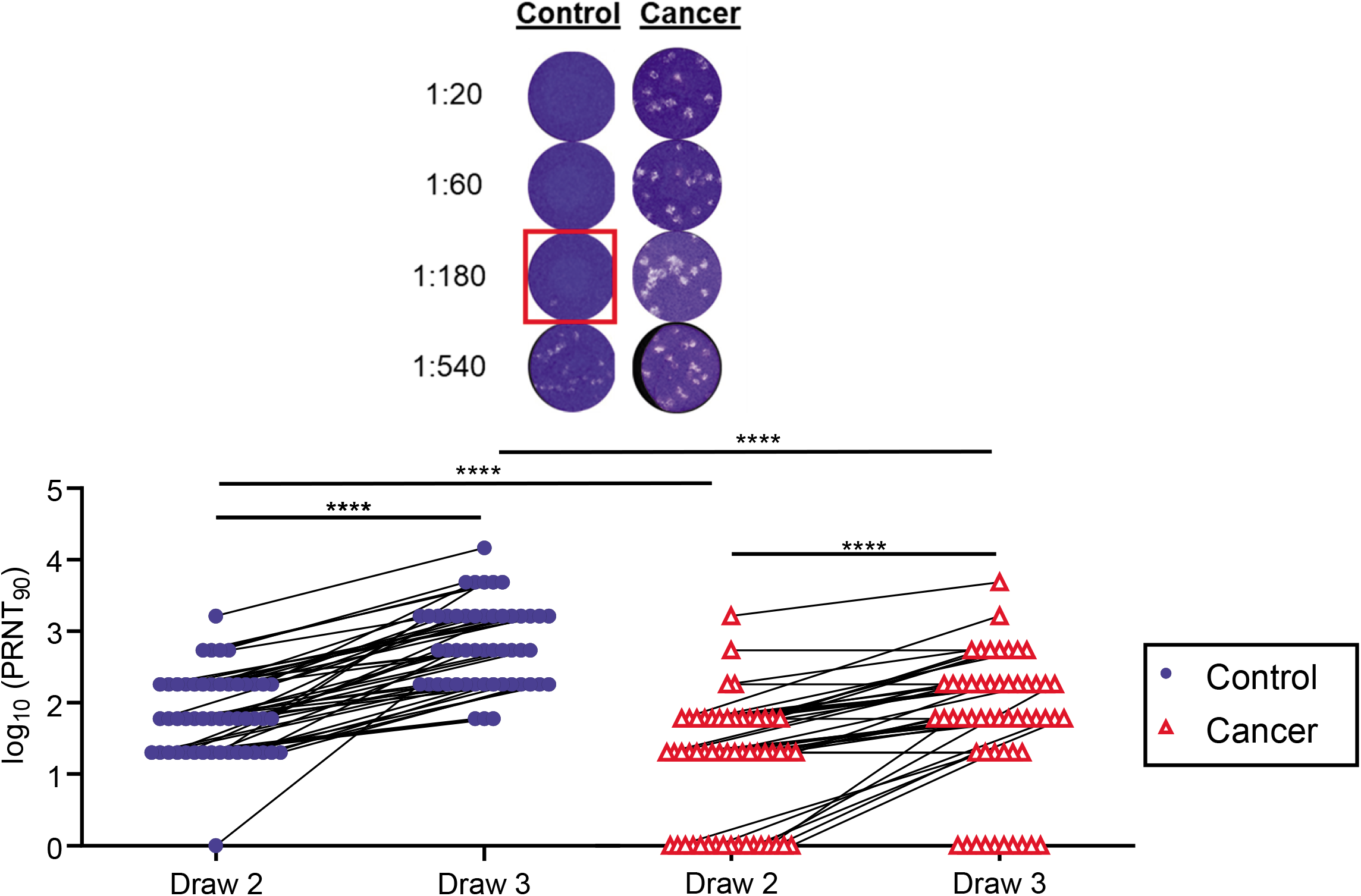
Neutralizing antibody responses of cancer and control cohorts to mRNA vaccination. Virus neutralization assays were performed using the WA1 isolate of SARS-CoV-2. Serial 1:3 dilutions of serum were performed and tested for the ability to prevent plaques on Vero cells. The lowest concentration capable of preventing >90% of plaques was considered to be the PRNT90 value. Example images are shown for the control and cancer cohorts with the red box indicating the PRNT90 titer. Quantification is shown below. Lines connect the same individual across timepoints. There is a statistically significant difference between draw 2 and draw 3 between cancer and control cohorts (p < 0.0001) and the average rate of change is increasing at a steeper rate in the control cohort (p = 0.0002). ****p<0.0001 by repeated measures ANOVA.

Prior studies have found that potentially protective T cell responses can be observed in COVID-19 convalescent individuals and in animal models when antibody levels are very low, such as after asymptomatic infections^38–41^. Moreover, the magnitude of T cell responses correlates relatively poorly with neutralizing antibody titers^42^. To quantify T cell responses in our healthy and cancer cohorts, peripheral blood mononuclear cells (PBMCs) were cultured overnight with either activating anti-CD3 antibodies (Extended Data Figure 4a) or a pool of overlapping Spike protein peptides capable of presentation on both HLA-I and HLA-II (Figure 3a). ELISPOT assays were then performed to quantify interferon gamma (IFN*γ*)-producing T cells relative to paired control wells in which no peptides were added. In the control cohort, we observed a marked increase in the median frequency of IFN*γ*+ T cells after the first vaccination relative to pre-vaccination timepoints (2.9 fold, p<0.0001), and a further increase after the second vaccination (2.6 fold, p<0.0001, Figure 3a). Within the cancer cohort, the first vaccination did not induce a statistically significant increase in the median frequency of Spike-specific IFN*γ*+ T cells at draw 2, but a clear increase was observed at draw 3 (4-fold, p<0.001, Figure 3a), though there was substantial variability in the response. Accordingly, T cell frequencies were reduced in the cancer cohort relative to healthy controls after the first vaccination and approached but remained below the levels observed in the control cohort after the second vaccination (p<0.05, Figure 3a). The majority of these responses likely reflect Spike peptide-specific CD4+ T cells^43, 44^, though CD8+ T cells may also contribute^45–47^. We re-tested samples with the highest Spike-specific T cell frequencies in the presence or absence of blocking antibodies against HLA-I and/or HLA-II to estimate CD8+ and CD4+ T cell responses, respectively. Substantial variation was observed across individuals in both the control and cancer cohorts, especially in HLA-I-dependent CD8+ T cell responses (Extended Data Figure 4b). Nonetheless, the data indicate that most individuals mount CD4+ and/or CD8+ responses.

**Figure 3:**
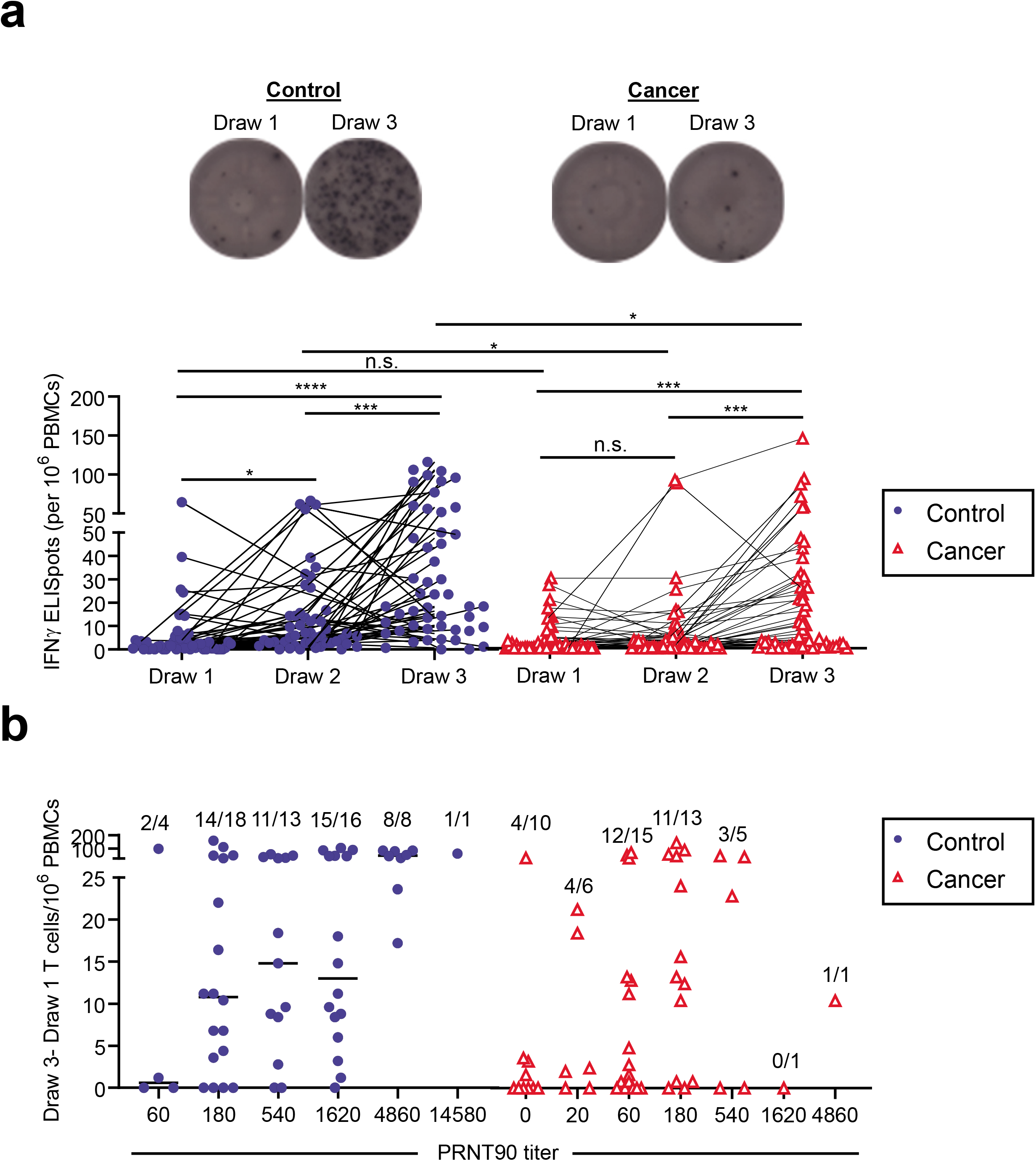
Spike-specific T cell responses of cancer and control cohorts to mRNA vaccination. **a,** PBMCs were cultured for 24 h in the presence or absence of a pool of overlapping Spike protein peptides. IFN*γ*-producing cells were quantified by ELISPOT. Example images are shown for the control and cancer cohorts at timepoints 1 and 3. Quantification is shown below of the no peptide background-subtracted data. Lines connect the same individual across timepoints. There is a statistically significant difference in slopes between cancer and control cohorts (p = 0.0284) and the average rate of change is increasing steeper in the control cohort. While overall (draw 1 to draw 3, p = 0.0455) the rates of change between draw 1 to draw 2 and draw 2 to draw 3 were not statistically significant (p-values = 0.0642 and 0.9891, respectively). The inability to detect a statistical difference, particularly between draw 1 and draw 2, is likely due to sample size and variability as the cancer cohort difference is flatter than the cancer cohort between these two draws. **b,** Draw 1 Spike-specific T cell frequencies were subtracted from draw 3 frequencies as calculated in **a** and plotted by PRNT90 titers. Frequencies of individuals with detectable Spike-specific T cells are shown above each group; analyses were done on the log-transformed scale. There was a statistically significant difference in slopes between the cancer and control cohorts (p = 0.0284). The primary difference in rates between the two cohorts were between draw 1 and draw 3 (p = 0.0455), the differences between draw 1 and draw 2 and draw 2 and draw 3 were not statistically significantly different (p = 0.0642 and 0.9891, respectively). *p<0.05; ***p<0.001 by repeated measures ANOVA.

To determine whether participants with poor neutralizing antibody titers might be partially protected by T cell responses, we examined T cell frequencies grouped by neutralizing antibody titers. Spike protein peptide-specific T cell frequencies at draw 1 were subtracted from the final draw 3 numbers to define individuals who mounted a response to vaccination. As has previously been described in post-infection responses^38, 42^, Spike protein peptide-specific T cell frequencies correlated relatively poorly with neutralizing antibody titers for both the healthy and cancer cohorts (Figure 3b). These data revealed that 4/10 cancer patients had detectable T cell responses even when PRNT90 titers were undetectable (Figure 3b). These data demonstrate that despite chemotherapy-induced immune suppression, relatively few cancer patients failed to make any detectable neutralizing antibody or T cell response. Nonetheless, these responses were substantially diminished relative to the control cohort, likely due to anti-cancer therapy.

One drawback to these interpretations is that the median age of the cancer cohort was greater than that of controls (Table 1). This raises concerns that some of the differences we observed were effects of age rather than of anti-cancer therapy. Yet the only immunological parameter that showed an age-dependent effect was anti-RBD antibody levels, which did show a decline with increasing age in the control cohort (p_interaction_ = 0.01, Extended Data Figure 5a). However, we observed no such age-dependent differences in the cancer cohort. Moreover, no other immunological parameters such as neutralizing antibody levels or T cell responses were altered as a function of age (Extended Data Figure 5b-c). Furthermore, when limiting the data to participants > 39 years of age (upper three quartiles), the differences between the two cohorts remained statistically significant for all immunological parameters (p-values < 0.0001). These data are consistent with the only modest age-dependent effects in immune responses reported by Pfizer/BioNTech^43^.

Because we assessed responses between 5-11 days after the second immunization, we examined whether there were any time-dependent changes in responses within this window. RBD-specific antibodies, neutralizing titers, and T cell responses did not obviously differ as a function of time after vaccination (Extended Data Figure 6a). Most control and cancer cohort participants were tested at 7-8 days after vaccination (Extended Data Figure 6a). Though the study was not powered for subgroup analyses, we also examined whether the tumor subtype might influence immune responses. No obvious differences were observed in antibody and T cell responses between breast, pancreatic, and other tumor types (Extended Data Figure 6b). We noted that one participant in the cancer cohort mounted a much stronger antibody response than the rest of the group (Extended Data Figure 6a-b). This participant had self-reported prior COVID-19 infection, despite seronegativity prior to the first immunization. Yet three other participants with self-reported prior COVID-19 showed no unusual patterns of antibody or T cell responses (Extended Data Figure 6c). Based on our initial inclusion criteria of seronegativity prior to vaccination, we retained these individuals in our analyses, but note that at least one of these four participants might be mounting recall, rather than primary responses. Together, these data suggest that anti-cancer therapy hampers immune responses to the Pfizer/BioNTech mRNA vaccines. The effectiveness of these diminished immune responses in preventing COVID-19 is difficult to predict.

Memory B cell frequencies are predictive of anamnestic responses following booster vaccination^24^, and presumably viral exposures. We first quantified RBD-specific CD19+ B cells in the control and cancer cohorts (Figure 4a) using antigen tetramers. We also simultaneously measured memory B cells that bound the S1 region of spike protein but not RBD (Figure 4a), as the N-terminal domain of S1 contains several neutralizing epitopes^48^. Within the control cohort, a clear increase in total RBD-specific B cells was observed after vaccination, but despite a trend, no statistically significant increase was observed in such cells in the cancer cohort (Extended Data Figure 7). Neither the control nor the cancer cohort showed a significant increase in S1-specific B cells with vaccination (Extended Data Figure 7).

**Figure 4:**
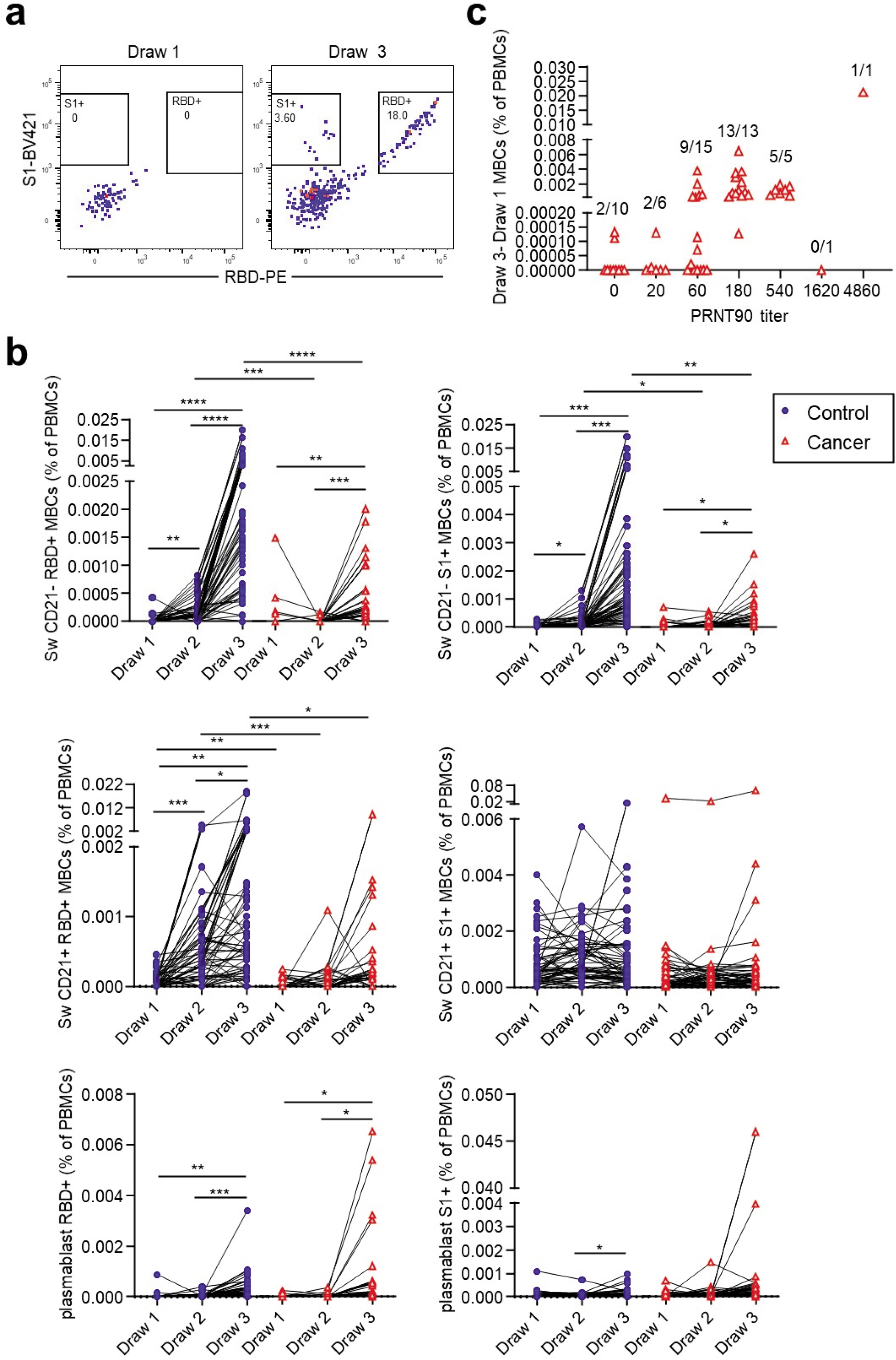
Memory B cell responses of cancer and control cohorts to mRNA vaccination. **a,** Example gating strategy of RBD- and S1-specific CD21-isotype-switched memory B cells (full gating strategy is shown in Extended Data Figure 1). **b,** Quantification of memory B cell and plasmablast subsets after vaccination. Isotype-switched (Sw) memory B cells expressing or lacking CD21 are shown in plots along with plasmablasts. Cells that bind both RBD and S1 are annotated as RBD+, whereas cells that are specific only for S1 are denoted as S1+. Lines connect the same individual across blood draws, analyses were done on the arcsin of the square-root transformation, to standardize the small percentages. There is a statistically significant difference in slopes between cancer and control cohorts for RBD+ and S1+ (p < 0.0001 and < 0.0001, respectively) and the average rate of change is increasing in the control cohort for both RBD+ and S1+, though the magnitude is only statistically significant between draw 2 and 3 for RBD+ between the cancer cohort and the control cohort (p=0.0991 and p< 0.0001, respectively) and S1+ (p = 0.3074 and < 0.0001, respectively). **c,** RBD-specific DN2, DN3, and S1- and RBD-specific isotype-switched CD21-memory B cells were added for the cancer cohort. Summed draw 1 memory B cell frequencies were subtracted from the summation of draw 3 frequencies for each individual. These values were grouped by PRNT90 titers. The average rate of change was statistically significantly different for RBD+ (p < 0.0001), with an increasing trend in the control cohort, and difference in rates between draw 1 and draw 2, and not between draw 2 and draw 3 (p = 0.0160 and 0.1059, respectfully). There was no statistically significant difference in slopes between cancer and control cohorts for S1+ (p=0.2239). Frequencies of individuals with detectable memory B cells are shown above each group. *p<0.05; **p<0.01; ***p<0.001; ****p<0.0001 by repeated measures ANOVA.

To gain more resolution, we examined antigen-specific frequencies within defined memory B cell subsets. These subsets exhibit different behaviors in recall responses, generating either plasmablasts or new germinal centers^49–54^. These lineage potentials correlate with antibody isotype and other markers^49–54^. We therefore quantified RBD- and S1-specific naive B cells, plasmablasts, and memory B cell subsets after vaccination. These subsets include IgG+ and IgM+ CD27+ CD21+ classical resting memory B cells^54^, CD27+ CD21- CD11c+ pre-plasmablast memory B cells^54–56^, CD27-IgD- CD11c- CD21+ DN1 cells, CD27-IgD- CD11c+ DN2 cells, and CD27-IgD- CD11c- CD21-DN3 cells (Figure 4a, Extended Data Figure 1). In the control cohort, we observed a clear increase in the frequency of isotype-switched RBD-binding CD21+ classical resting memory B cells as well as CD21-pre-plasmablast memory B cells after each vaccination (Figure 4b). Isotype-switched S1-binding CD21-memory B cells were also observed to increase after each vaccination of the control cohort (Figure 4b). Within the cancer cohort, we also observed isotype-switched RBD- and other S1-specific pre-plasmablast CD21-memory B cells, but only after the second immunization, and these levels were ∼10-fold lower than those observed in the control cohort (Figure 4b). We were unable to detect isotype-switched RBD-or S1-specific classical resting memory B cells above pre-vaccination levels in the cancer cohort (Figure 4b). Some other RBD- and S1-binding memory B cell subsets were detectable in the healthy and cancer cohorts, but in general the frequencies of these cells were substantially lower than the isotype-switched CD27+ subsets and not consistently increased with each immunization (Extended Data Figure 8). We were unable to detect antigen-specific cells above background levels in naive B cells (Extended Data Figure 8). Thus, RBD- and S1-specific cells early after vaccination are enriched in IgG+ memory subsets. These cells are biased towards plasma cell fates^52, 54^, though secondary germinal centers could conceivably arise from classical CD21+ memory cells^53^. In both cohorts, we observed increases in RBD-specific antibody-secreting plasmablasts, with no statistically significant differences between cohorts (Figure 4b). S1-specific plasmablasts were not readily apparent in either group (Figure 4b). This might be partly due to poor survival of plasmablasts after freezing and in part due to lower surface B cell receptor levels^57^.

We next examined whether RBD- and S1-specific memory B cells could be detected in cancer patients with no or low levels of neutralizing antibodies. Prior studies have shown that memory B cell numbers and specificities correlate only modestly with serum antibodies^58–62^. CD21-RBD+ and S1-specific memory B cell frequencies at draw 3 were added to DN2 and DN3 RBD+ memory B cell frequencies for each cancer patient, as these subsets were the only ones in which cancer patients consistently showed vaccine-induced increases (Figure 4b and Extended Data Figure 8). Next, the corresponding pre-vaccination draw 1 frequencies were subtracted to correct for background levels in each patient. These net memory B cell frequencies were then plotted as a function of virus neutralization titers. Patients without detectable neutralizing antibodies also generally lacked RBD- and S1-specific memory B cells (Figure 4c). In contrast, patients with modest but detectable neutralizing antibody titers consistently showed RBD- and S1-specific memory B cells after the second immunization (Figure 4c). These data suggest that patients with low but detectable Spike-specific antibodies would likely generate anamnestic responses after a third immunization, conceivably approaching levels seen in healthy controls after the second vaccination.

To directly determine whether and how immunity can be improved by a third vaccination, we initiated an interventional trial for our cancer cohort. Twenty of the original cohort agreed to participate and met the inclusion criteria. There were no statistically significant associations between participation and draw 3 RBD-specific antibodies, neutralization titers, or Spike-specific T cells. However, we note the study was not powered sufficiently to specifically preclude these differences. All patients were contacted within 2–4-week windows for adverse events. There were no Serious Adverse Events (SAEs) noted (Table 2), with 9 (45%) participants experiencing injection site pain. Other minor Adverse Events (AEs) included: generalized myalgia (15%); bone pain (5%); fatigue (10%); chills (10%); and appetite loss (5%). There were no obvious demographic differences between the 20 participants and the original cancer cohort; however, these participants did have a shorter window between administration of cancer treatment and blood draws for analyses. Patients in this cohort had gastrointestinal cancers predominantly (75%) compared with 51% in the original cancer cohort; the remaining five participants (25%) had a breast cancer diagnosis compared to 42% in the original cancer cohort.

**Table 2:** Adverse Events.

| **Adverse Event<sup>£</sup>** | **N (%)** |
| --- | --- |
| Appetite Loss | 1 (5%) |
| Chills | 2 (10%) |
| Fatigue | 2 (10%) |
| Generalized Bone Pain | 1 (5%) |
| Generalized Myalgias | 3 (15%) |
| Injection Site Myalgia | 9 (45%) |

<sup>£</sup>All were mild events

Blood samples were acquired at the time of the third shot (Draw 4, Extended Data Figure 9a) and 1 week afterwards (Draw 5). RBD-specific antibodies, virus-neutralizing antibodies, and Spike-specific T cells were quantified. A modest but consistent and statistically significant increase from Draw 4 to Draw 5 was observed in mean RBD-specific antibody titers 1 week after the third shot relative to the pre-boost levels (0.49 vs. 0.72, p=0.02, Figure 5a). This was accompanied by a 3-fold increase in median virus-neutralizing antibody titers (60 vs. 180, p<0.0001, Figure 5b). Interestingly, two participants who had no detectable neutralizing antibodies at Draw 3 showed an increase at Draw 4, even prior to the booster immunization (Figure 5b). In both cases, neutralizing antibodies increased further after the third shot (Figure 5b). We observed no overall increase in T cells after booster immunization of the cancer cohort (Figure 5c). Because participants received the third shot between 42-111 days after the third draw, we examined whether the duration of time between immunizations might influence the magnitude of the antibody response, as has been reported for doses 1 and 2^63^. Yet we observed no correlation between the time between doses and the magnitude of the RBD-specific or neutralizing antibody recall responses (Extended Data Figures 9b-c).

**Figure 5:**
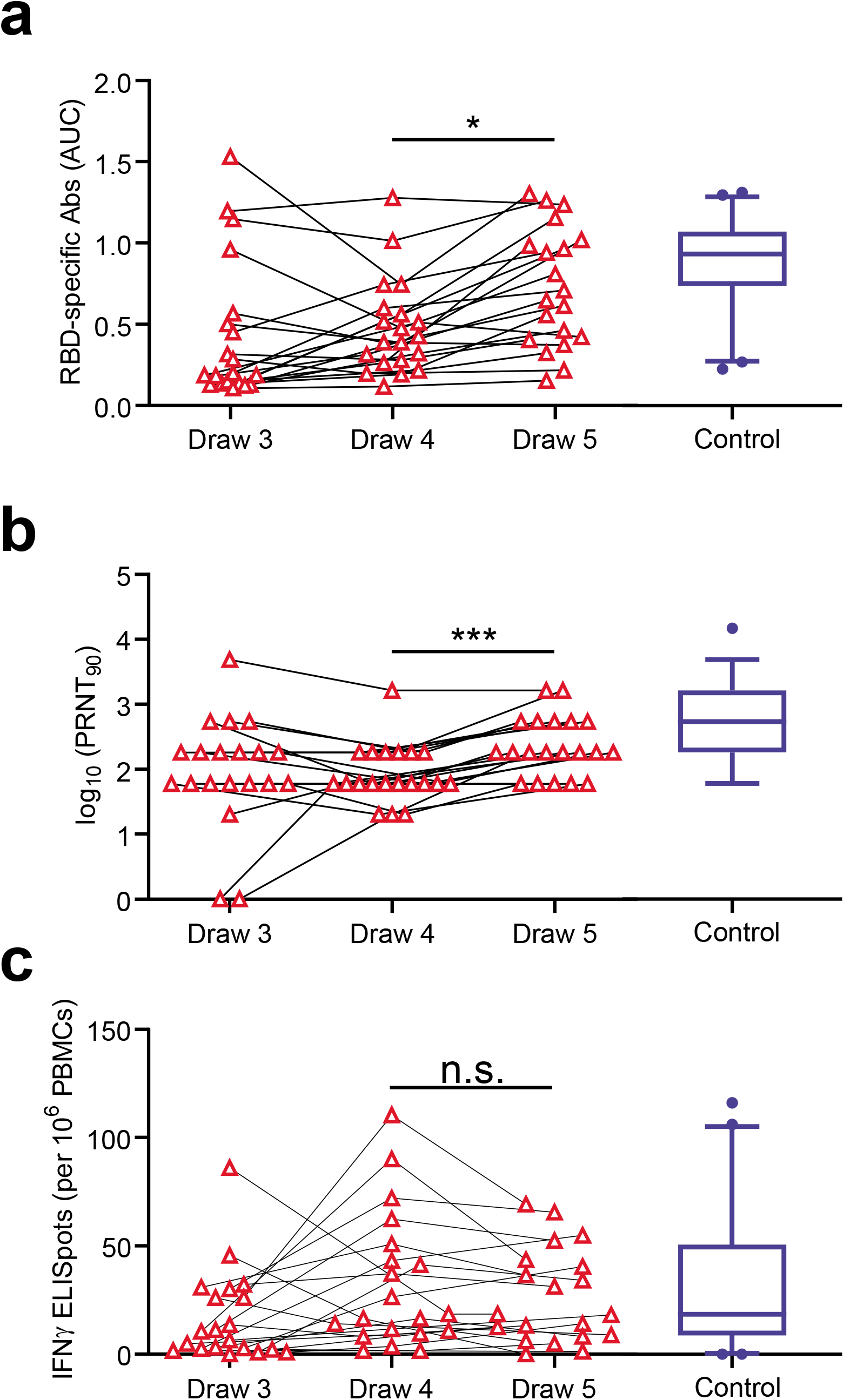
Antibody responses improve after a third immunization. **a**, RBD-specific antibody titers were quantified at the time of the 3rd immunization (draw 4) and 1 week afterwards (draw 5). Data from draw 3 are the same as those in Figure 1d and are shown again for context. Blue boxplot shows 95% confidence intervals for the control cohort at draw 3. **b,** Neutralizing antibody titers were quantified at the time of the 3rd immunization (draw 4) and 1 week afterwards (draw 5). Data from draw 3 are the same as those in Figure 2. Blue boxplot shows 95% confidence intervals for the control cohort at draw 3. **c,** Spike-specific T cell responses were quantified at the time of the 3rd immunization (draw 4) and 1 week afterwards (draw 5). Data from draw 3 are the same as those in Figure 3a. Blue boxplot shows 95% confidence intervals for the control cohort at draw 3. P-values were calculated using a paired t-test. Statistics are shown only for draw 4 vs. draw 5 comparisons.

Prior studies of vaccinations of COVID-19 convalescent individuals revealed a strong correlation between pre-existing memory B cells and the magnitude of the antibody response after immunization^24^. To determine whether such a relationship could be observed in our cancer cohort, we plotted the draw 3 RBD-specific memory B cell frequencies (switched CD21-, switched CD21+, DN2, and DN3) against the change in RBD-specific antibodies after the booster immunization. Unexpectedly, we observed no correlation between these parameters (Extended Data Figure 9d). A similar lack of correlation was observed between summed RBD and S1-specific memory B cells and boosted virus-neutralizing antibody titers (Extended Data Figure 9e). These data suggest that the presence of memory B cells in the cancer cohort may be a reasonable indicator that an antibody recall response will occur after the third immunization, but unlike in healthy individuals^24^, memory B cell frequencies are not quantitatively predictive of the magnitude of the response.

To begin to explain this lack of correlation between memory B cells and subsequent anamnestic responses, we performed comparisons of memory B cell frequencies, antibody levels, and T cell responses prior to the third shot at draw 3 (Extended Data Figure 10). Most parameters were well-correlated with each other in the control cohort in biologically rational ways. For example, isotype-switched CD27+ and DN2 subsets clustered together and were highly correlated (Extended Data Figure 10). Within the cancer cohort, however, memory B cell subsets were not well-correlated and did not cluster with each other (Extended Data Figure 10). This implies a lack of coordination between aspects of the response that are normally linked, which in turn may lead to quantitatively unpredictable recall responses.

## Discussion

The COVID-19 pandemic has dramatically affected the world, with a profound impact on cancer patients and their care. With high rates of transmission, even mitigation strategies have not been enough to decrease mortality rates from COVID-19 in patients with active cancer. Thus, the development of mRNA vaccines directed against SARS-CoV-2 was anxiously awaited in the cancer community. Given that the Pfizer/BioNTech trials did not include patients with active malignancies^3^, the efficacy of these vaccines in solid tumor patients on active therapy was not reported. While prior studies in patients with colorectal and breast cancers on active chemotherapy who receive influenza vaccination show that patients can mount a serological response, the immunogenicity of the mRNA COVID vaccines in these patients is largely unknown^64, 65^. A recent study in *JAMA* looked at 658 organ transplant recipients and demonstrated a lower antibody response after both doses of the mRNA COVID-19 vaccines when compared to immunocompetent participants^21^. Similarly, early studies suggest that cancer patients do not mount the same antibody responses as healthy controls^22^.

Our results agree with certain aspects of these findings but differ with and extend upon several key points. As with a recent study on immunocompromised cancer patients^22^, we observed lower overall antibody and T cell responses compared with control cohorts. Yet in contrast to these findings, we observed that the majority of patients seroconverted after the first immunization, as measured by live virus neutralization assays. This frequency further increased after the second vaccination. These differences could potentially be explained by the nature of our cohort, which did not include patients on immunotherapy or patients with hematologic malignancies. In addition, neutralization assays using authentic viral isolates, as we used here, tend to be more sensitive than experiments performed with Spike protein-pseudotyped lentiviruses^34, 35^. The ability to detect low levels of neutralizing antibodies is important when interpreting the potential value of vaccinating immunocompromised individuals.

There are several lines of evidence to suggest that the threshold for protection against COVID-19 may be relatively low. First, following natural infection, the levels of neutralizing antibodies are often quite low but symptomatic re-infections are rare^66–70^. Second, despite modest induction of overall immune responses, a single dose of mRNA vaccine provides reasonable protection against COVID-19^3–5^. Third, non-human primate studies have shown that low levels of passively transferred antibodies are protective against large infectious doses of SARS-CoV-2^41^. In these models, even when antibodies drop below protective levels, T cells can compensate and cooperate with residual antibodies to confer protection^41^. T cell responses are likely protective against severe disease in humans as well^38, 39, 71^. In this sense, it is encouraging that we also observed T cell responses in the majority of vaccinated cancer patients, including nearly half that mounted undetectable neutralizing antibody responses. Unlike antibodies, these T cell responses were only modestly reduced relative to the control cohort. Of note, many individuals possess Spike-reactive memory T cells, but not B cells or antibodies, even prior to SARS-CoV-2 exposure or vaccination^72^. It is possible that these pre-existing coronavirus cross-reactive memory T cells dominate after vaccination, diminishing the negative impact of anti-cancer therapy on immunization. The resulting CD4+ T cells could potentially help naive B cells participate in subsequent responses to vaccines and infections, which may help explain the poor correlations we observed between memory B cell frequencies and the magnitude of the recall responses in cancer patients. Given that T cells reduce viral loads and disease even when neutralizing antibody levels are low^38, 40, 41, 73^, these data suggest that vaccination will confer at least partial protection and reduce the likelihood of severe COVID-19 in most cancer patients.

Nonetheless, when compared with individuals not on immunosuppressive therapy, the magnitudes of vaccine-induced antibody and T cell responses were substantially reduced in cancer patients. These reduced levels may be particularly problematic when faced with variants possessing some neutralizing antibody-evading mutations, such as beta, gamma, or delta^74, 75^. Some participants in our cohort failed to mount detectable antibody or T cell responses by 1 week post-immunization, although several did show improvement over time. This seems likely to diminish vaccine effectiveness relative to the benchmark of 94-95% in non-immunocompromised populations^3, 4^. Several recent studies have reported improved antibody responses in transplant recipients after a third dose, though neutralizing antibodies and T cells were not quantified^76, 77^. We therefore initiated a trial to determine whether a third immunization would improve immunity in our cohort on active anti-cancer therapy. Interestingly, two participants who initially failed to mount detectable responses by 1 week after the second vaccination later displayed detectable antibodies prior to the third dose; one patient with cholangiocarcinoma on Gemcitabine/Cisplatin/nab-paclitaxel and another pancreatic cancer on Gemcitabine/Cisplatin. This suggests that for at least a subset of the non-responding cancer cohort, antibody responses may be delayed but not completely absent. After the third immunization, neutralizing antibody levels improved in 16 out of 20 participants. In all but one of the participants who improved, neutralizing antibody titers reached 180 or greater. In non-human primate and modeling studies, this level is protective against disease^41, 78^, though new variants may change these considerations. Nonetheless, the overall antibody increases induced by the booster immunization were fairly modest, and, for reasons that are unclear, no further improvement was observed of circulating Spike-specific T cell frequencies. Of the 4 participants who did not improve antibody levels after the third vaccination, one had very high antibody levels already after the first dose and had reported prior COVID-19 in a questionnaire, despite being seronegative at the outset of the study. The lack of impact of immunization on those with high starting antibody levels has been noted in other studies of COVID-19 convalescent individuals^24, 79^, perhaps because these pre-existing serum antibodies mask antigens and prevent additional B cell responses^80^.

Our cancer cohort naturally had an expected heterogeneity in terms of cancer diagnoses, the types of cytotoxic therapy patients received, and the timing of these therapies relative to vaccine dose. Thus, it is difficult to draw conclusions related to which solid tumors were associated with a better vaccine response or which therapies correlated with the non-responders. Yet it is worth noting that most of the initial non-responders had blood collected for immune analysis 7-14 days after their most recent treatment with cytotoxic agents. This time course is aligned with a nadir in blood counts and the peak of myelosuppression from traditional chemotherapy agents. While the numbers are too small to draw strong conclusions, these findings are certainly hypothesis-generating and merit further exploration to better understand the ideal timing for vaccination in patients on active immunosuppressive therapy. Our cancer cohort was also on average older than participants in the control cohort. There did appear to be an age-moderated effect within the control group on anti-RBD titers, which in turn could impact the magnitude of the differences we observed between the control and cancer cohorts. Yet no other immunological parameters were similarly affected and we did not observe age-moderated effects within the cancer cohort for any immunological parameter. Thus, the major driver of diminished responses in the cancer cohort is likely to be anti-cancer therapy rather than age.

Together, our data suggest that most cancer patients on active chemotherapy are likely to improve antibody levels and protection from COVID-19 after a third immunization. Yet given the relatively modest increases in antibodies and recalcitrance of T cells, expectations should remain tempered as to the degree of benefit. Quantitative antibody tests can potentially be used to select individuals who need and would most benefit from a booster.

## Methods

### Participant Selection

This protocol was approved by the University of Arizona Institutional Review Board and activated in January 2021. Participants were recruited to the control cohort during the Phase 1B Pima County COVID-19 vaccine rollout. Participants scheduled for vaccine appointments at Banner University Medical Center North site were approached with the IRB approved consent and sequentially enrolled. Thereafter, patients with cancer diagnosis were enrolled at the University of Arizona Cancer Center. Informed consent was obtained for all participants. Eligible solid tumor cancer patients had to have active disease and be receiving ongoing cytotoxic systemic therapy. Patients receiving immunotherapy were excluded. Demographic information was collected in addition to cancer diagnosis and type of anti-cancer therapy. Dates of last treatment prior to vaccine administration were also noted. In total, 73 control cohort participants were consented and 65 completed all three blood draws and both vaccine shots; five did not come in for their blood draw and eleven did not show up for their scheduled vaccinations. Fifty-six cancer patients were consented for the study and 53 completed all three blood draws and received both shots. All of the cancer cohort participants received the Pfizer Vaccine, 61 enrolled participants in the control cohort received the Pfizer vaccine and 12 received the Moderna vaccine. For consistency, analyses are restricted to those participants that received the Pfizer vaccine. There were four control cohort participants that were seropositive based on the University of Arizona COVID-19 ELISA pan-Ig Antibody Test; all of these participants were removed from analyses. The complete study sample size is 53 cancer cohort patients and 50 control cohort participants. For the interventional booster all 53 patients in the cancer cohort were considered for continued eligibility and re-consenting. Reasons for drop-out from the observational cancer cohort included eleven participants ineligible due to going off chemotherapy (11.3%); ten died (unrelated to vaccine) or were in hospice (19%); six had health or safety concerns per investigator (11.3%); and eleven declined to participate (11.3%). The full CONSORT flow diagram is shown in Extended Data Figure 11. Blood samples were drawn for analysis prior to the administration of a third booster dose of the Pfizer mRNA vaccine. A final blood draw was performed on all 20 patients between 5-11 days from the time of the 3rd vaccine. Patients in this cohort were contacted at 2 weeks (+/- 3 days) and 4 weeks (+/- 7 days) post booster dose for adverse events and serious adverse event monitoring.

### Peripheral blood mononuclear cell and serum preparation

Twenty mL of blood was collected by venipuncture in heparinized Vacutainer tubes (BD) and an additional 10 mL was collected in clot activating non-heparinized Vacutainer tubes. After >30 minutes at room temperature, non-heparinized tubes were spun at 1200 x g for 10 minutes, and serum was collected and frozen in 1 mL aliquots at −20°C. For peripheral blood mononuclear cells (PBMCs), 15 mL of Ficoll-Paque Plus (Fisher Scientific) was added to 50 ml Leucosep tubes (Greiner) and spun for 1 minute at 1000 x g to transfer the density gradient below the filter. Twenty mL of blood from the heparinized tubes were then poured into the top of the Leucosep tube and then spun at 1000 x g for 10 minutes at room temperature with the brake off. The top plasma layer was carefully collected and frozen at −20°C and the remaining supernatant containing PBMCs above the filter was poured into a new 50 mL conical tube containing 10 ml phosphate buffered saline (PBS) and spun at 250 x g for 10 minutes. Cell pellets were resuspended in RPMI media containing 10% fetal calf serum and counted on a ViCell XR (Beckman Coulter). Cells were resuspended to a concentration of 2 x 10⁷ cells/mL in RPMI media containing 10% fetal calf serum. An equal volume of 80% fetal calf serum + 20% dimethyl sulfoxide was added dropwise and inverted once to mix. Suspensions were distributed at 1 mL/cryovial and frozen overnight at −80°C in Mr. Frosty freezing chambers (Nalgene). Vials were then transferred to storage in liquid nitrogen.

### ELISAs and quantification of antibody titers

Serological assays were performed as previously described. RBD was purchased from GenScript (catalog # Z03483) and S2 subdomain of the SARS-CoV-2 S glycoprotein was purchased from Sino Biological (catalog # 40590-V08B). To obtain titers and single-dilution OD450 values, antigens were immobilized on high-adsorbency 384-well plates at 5 ng/ml. Plates were blocked with 1% non-fat dehydrated milk extract (Santa Cruz Biotechnology #sc-2325) in sterile PBS (Fisher Scientific Hyclone PBS #SH2035,) for 1 h, washed with PBS containing 0.05% Tween-20, and overlaid for 60 min with either a single 1:40 dilution or 5 serial 1:4 dilutions beginning at a 1:80 dilution of serum. Plates were then washed and incubated for 1hr in 1% PBS and milk containing anti-human Pan-Ig HRP conjugated antibody (Jackson ImmunoResearch catalog 109-035-064) at a concentration of 1:2000 for 1 h. Plates were washed with PBS-Tween solution followed by PBS wash. To develop, plates were incubated in tetramethylbenzidine prior to quenching with 2N H2SO4. Plates were read for 450nm absorbance on CLARIOstar Plus from BMG Labtech. All samples were also read at 630nm to detect any incomplete quenching. Any samples above background 630nm values were re-run. Area Under the Curve values were calculated in GraphPad Prism (v9).

### T cell assays

Frozen PBMCs were thawed by mixing with 10 mL of pre-warmed RPMI 1640 media (Gibco) containing 10% fetal calf serum (Peak Serum #PS-FB1), 1X Penicillin-Streptomycin (HyClone #SV30010) and 0.03 mg/mL DNAse (Sigma #DN25-100) in a 15 mL conical tube and spun at 1650 rpm for 5 minutes. Cell pellets were resuspended in 1 mL of X-VIVO 15 media with Gentamicin and Phenol Red (VWR #12001-988) containing 5% male human AB serum (Sigma #H4522-100ML), and incubated in 24-well plates overnight at 37°C with 5% CO2. 250 μL of each sample was plated on a 96-well round bottom plate and spun at 1650 rpm for 3 minutes, and then resuspended in 150 μl X-VIVO 15 media with 5% male human AB serum containing either 0.6 nmol PepTivator SARS-CoV-2 Prot_S peptide pool (Miltenyi Biotec #130-126-701) for antigen specific T cell stimulation, or positive control anti-CD3 mAb CD3-2 from Human IFN-γ ELISpot plus kit (Mabtech #3420-4APT-2), or blank media as negative control. In some experiments, 10 μg/ml blocking antibodies against HLA-I (W6/32, Biolegend) and/or HLA-II (Tü39, Biolegend) were included. Cell suspensions were transferred to pre-coated IFN-γ ELISpot plates and incubated overnight at 37°C with 5% CO2. Plates were emptied, washed 5 times with 200 μl/well of PBS (Fisher Scientific Hyclone PBS #SH2035), and incubated for 2 hours at room temperature with 100 μl/well PBS containing 0.5% fetal calf serum and 1 μg/ml detection antibody (7-B6-1-biotin). Plates were washed as above and incubated for 1 hour at room temperature with 100 μl/well of PBS-0.5% FCS with 1:1000 diluted Streptavidin-ALP. Plates were washed as above and developed for 10-15 minutes with 100 μl/well substrate solution (BCIP/NBT-plus) until distinct spots emerged. Color development was stopped by washing extensively in tap water and left to dry. Spots were imaged and counted using an ImmunoSpot Versa (Cellular Technology Limited, Cleveland, OH) plate reader.

### Virus neutralization assays

SARS Coronavirus 2, Isolate USA-WA1/2020 (BEI NR-52281) was passaged once on Vero (ATCC #CCL-81) cells at a multiplicity of infection of 0.01 for 48 hours. Supernatant and cell lysate were combined, subjected to a single freeze-thaw, and then centrifuged at 3000 rpm for 10 minutes to remove cell debris. Plaque reduction neutralization tests (PRNT) for SARS-CoV-2 were performed as previously described. Briefly, Vero cells (ATCC # CCL-81) were plated in 96-well tissue culture plates and grown overnight. Serial dilutions of serum samples were incubated with 100 plaque forming units of SARS-CoV-2 for 1 hour at 37°C. Plasma/serum dilutions plus virus were transferred to the cell plates and incubated for 2 hours at 37°C, 5% CO2 then overlaid with 1% methylcellulose. After 72 hours, plates were fixed with 10% Neutral Buffered Formalin for 30 minutes and stained with 1% crystal violet. Plaques were imaged using an ImmunoSpot Versa (Cellular Technology Limited, Cleveland, OH) plate reader. The most dilute serum concentration that led to 10 or fewer plaques was designated as the PRNT90 titer.

### Flow cytometry

One mL of pre-warmed fetal calf serum was added to a frozen cryovial of PBMCs which was then rapidly thawed in a 37°C water bath. Samples were poured into 15 mL conical tubes containing 5 mL pre-warmed RPMI + 10% fetal calf serum and spun at 250 x g for 5 minutes, room temperature. Supernatants were removed and pellets were washed once with 500 ul PBS containing 5% adult bovine serum and 0.1% sodium azide (staining buffer). Cell pellets were then resuspended in 200 μl staining buffer containing 1μl each of anti-IgM-FITC (Biolegend clone MHM-88), anti-IgD-PerCP-Cy5.5 (Biolegend clone IA6-2), anti-CD11c-Alexa700 (Biolegend clone Bu15), anti- CD13-PE-Cy7 (Biolegend clone WM15), anti-CD19-APC-efluor-780 (eBioscience clone HIB19), anti-CD21-PE-Dazzle (Biolegend clone Bu32), anti-CD27-BV510 (Biolegend clone M-T271), anti-CD38-APC (Biolegend clone HIT2), RBD-PE tetramer, and S1-BV421 tetramer. Tetramer reagents were assembled by mixing 100μg/ml C-terminal Avitagged RBD or S1 (AcroBiosystems) with 100μg/ml streptavidin-PE (eBiosciences) or streptavidin-BV421 (Biolegend), respectively, at a 5:1 molar ratio in which 1/10 the final volume of streptavidin was added every 5 minutes. S1 and RBD tetramers were validated by staining 293T cells as a negative control or 293T-hACE2-expressing cells (BEI Resources NR-52511) as a positive control. PBMC samples were stained for at least 20 minutes, washed, and filtered through 70 μm nylon mesh. Data were acquired on either a BD LSR2 or Fortessa flow cytometer. Data were analyzed using FlowJo software.

### Statistical methods

The primary statistical endpoint for the observational study was the change in antibody-mediated neutralization of authentic live SARS-CoV-2 PRNT90 titers from baseline to draw 3 between participants in the control cohort and cancer cohort. This primary endpoint, powered as a non-inferiority hypothesis, was whether vaccine-acquired PRNT90 titers were the same in immunocompromised patients compared to healthy individuals. These methods typically require estimating the outcome under a non-inferiority margin; however, this criterion is not necessary given the obvious superiority PRNT90 titers seen in healthy participants compared to cancer patients at draw 3. The primary endpoint for the booster (interventional) study was the paired difference, using a paired t-test statistic, between draw 4 (booster shot) and draw 5 (7 days post booster) with secondary analyses examining the paired difference in RBD titers (as AUC) and Total T cells. Secondary analyses for the interventional study included comparing differences between pairwise differences in slope and between blood draws, e.g. draw 1 to draw 2 and draw 2 to draw 3 between cohorts using repeated measures analysis of variance (ANOVA) that adjusts for the correlation within an individual by use of an exchangeable covariance matrix and F-test statistics. Additionally, analysis of covariance was used to evaluate whether age moderated the between cohorts draw 3 differences for the semi-quantitative 1:40 serum dilution ELISA for RBD and S2 spike proteins as well as RBD AUC and neutralizing Ab titers. The cancer cohort was older, on average, with a mean age of 64 years compared to 42 in the control cohort. The possible mediating effect of age on the immunologic response seen between the cohorts was evaluated in two ways. First, age was added as an interaction effect to draw 3 differences between cohorts, using linear models with age as a continuous variable. Additionally, since the primary age difference between cohorts was the lack of participants in the lowest age quartile (< 38 years of age), the draw 3 differences were tested after removing this age group from control cohort, thus removing the lower age bias that could have been introduced by including these participants; this resulted in a much smaller sample size in the control cohort (n = 23) and was performed using linear modeling with a two-sample, two-sided, t-test at the 0.05 level of significance. All analyses were performed in GraphPad Prism 9 and the R programming language version 4.0.5.

## Data Availability

Raw data and statistical analysis code are available upon request to the corresponding authors. Figures 1-5 have associated raw data. Data will be made available without restrictions unless linked to identifiers in patient health information.

## Acknowledgements

This work was supported by National Institutes of Health grants R01AI099108 and R01AI129945 (D.B.) and from University of Arizona funds. We thank Marion Pepper and Jason Netland (University of Washington) for technical assistance with tetramer stains.

## Competing interests

Sana Biotechnology has licensed intellectual property of D.B. and Washington University in St. Louis. D.B. is a co-founder of Clade Therapeutics. B.J.L. has a financial interest in Cofactor Genomics, Inc. and Iron Horse Dx. P.C. receives research funding from Pfizer, BioAtla, Zentalis, Genentech, Eli-Lilly, Phoenix Molecular Designs, Amgen, Radius Pharmaceuticals, Carrick Therapeutics, and Angiochem and served on advisory boards for Novartis, Eli Lilly, Zentalis, Astra-Zeneca, Amgen, Bayer, Asthenex, Prosigna, Heron, Puma Biotechnology and Oncosec. R.T.S. receives research funding from Merck, Rafael Pharmaceuticals, ImmunoVaccine, Bayer, SeaGen, Exelixis, Pieris, LOXO Oncology, Novocure, NuCana, QED and has served as a consultant/advisor to Merck, Servier, Astra-Zeneca, EMD Serono, Taiho, QED, Incyte, Genentech, Basilea.

### Data, Code, and Materials Availability

Raw data and statistical analysis code are available upon request to the corresponding authors. Figures 1-5 have associated raw data. Data will be made available without restrictions unless linked to identifiers in patient health information. Unique biological materials, including patient sera and cells, are of limited quantity. When possible and upon reasonable request, we may be able to share extra materials in small quantities.

---

**Extended Data Figure 1:**
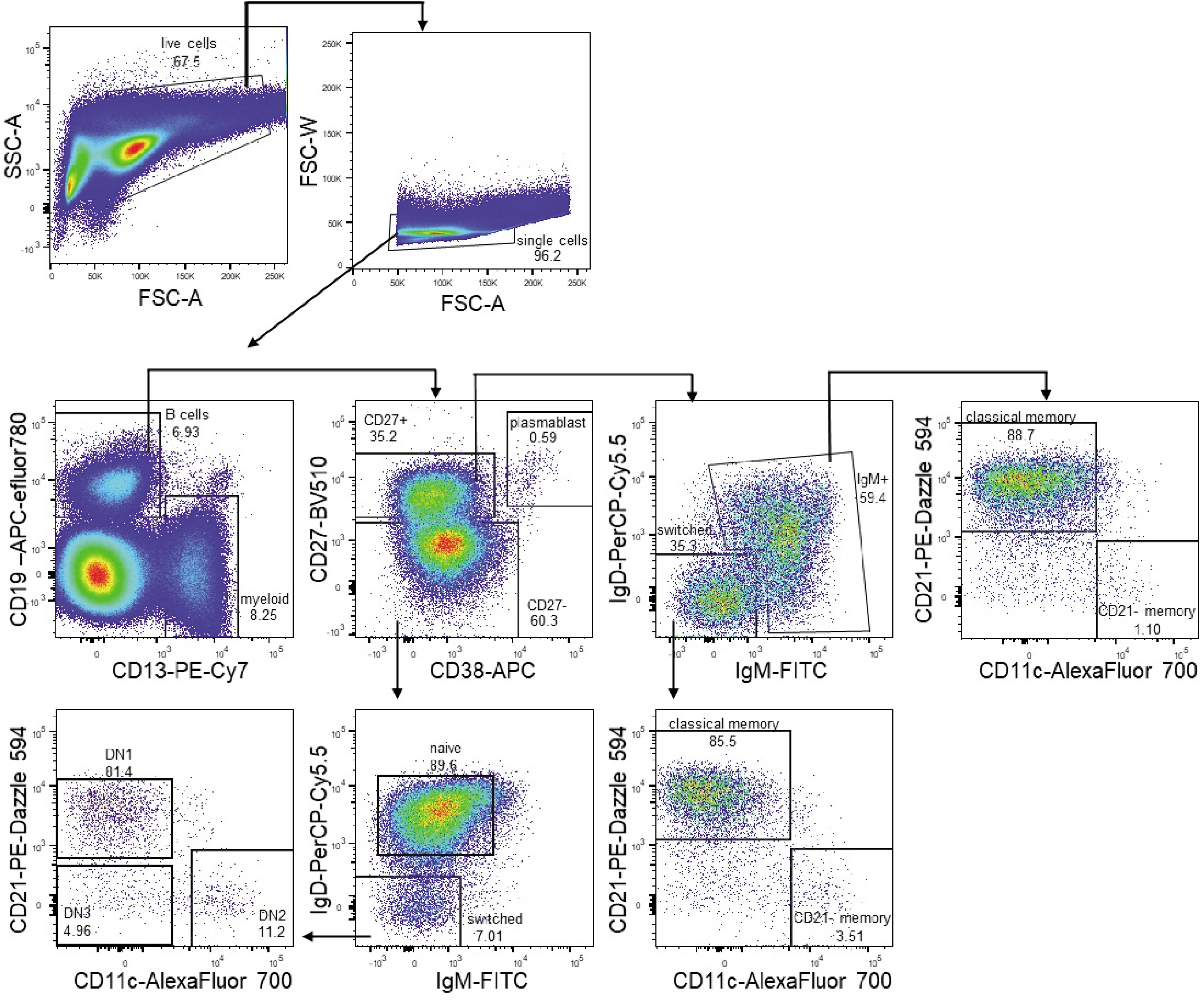
Gating strategy for B cells and myeloid lineages. Full gating strategy of naive and memory B cell subsets and plasmablasts (antigen-specific stains are shown in Figure 4a).

**Extended Data Figure 2:**
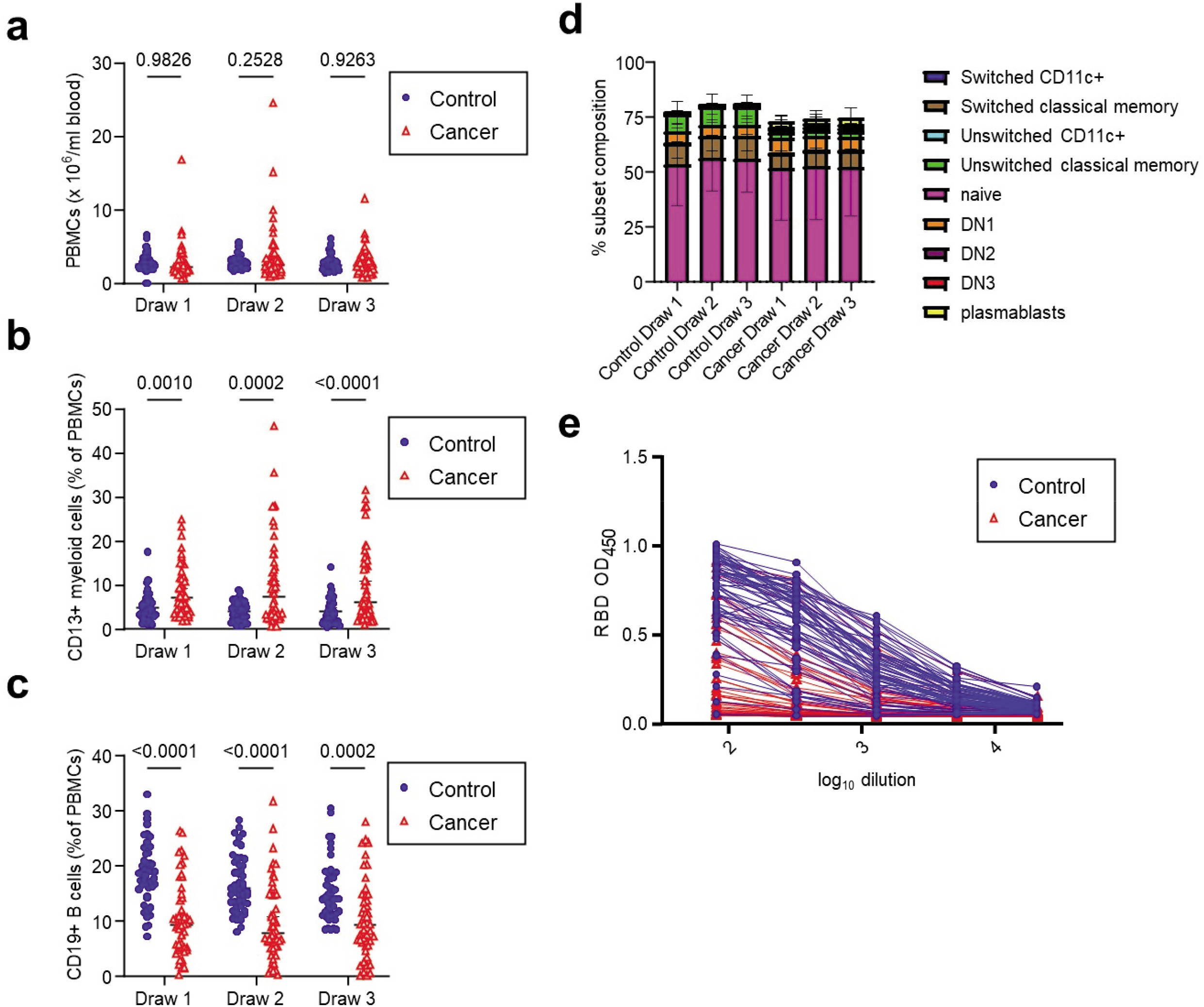
Cellular and serological characterization of blood samples from control and cancer cohorts. **a,** PBMC frequencies of blood samples at each timepoint. P-values were calculated by 2-way ANOVA in which individual samples across blood draws were paired. **b,** CD19+ B cell frequencies of blood samples at each timepoint. P-values were calculated by 2-way ANOVA in which individual samples across blood draws were paired. **c,** CD13+ myeloid cell frequencies of blood samples at each blood draw. P-values were calculated by 2-way ANOVA in which individual samples across draws were paired. **d,** B cell subset frequencies at each draw. **e,** Raw ELISA data for quantification of RBD titers shown in Figure 1d. A serum concentration beginning at 1:80 was serially diluted and area under the curve (AUC) values calculated. Lines connect the same individual at each dilution. Data from the third blood draw are shown for both the control and cancer cohort. *p<0.05; **p<0.01; ***p<0.001; ****p<0.0001 by 2-way ANOVA.

**Extended Data Figure 3:**
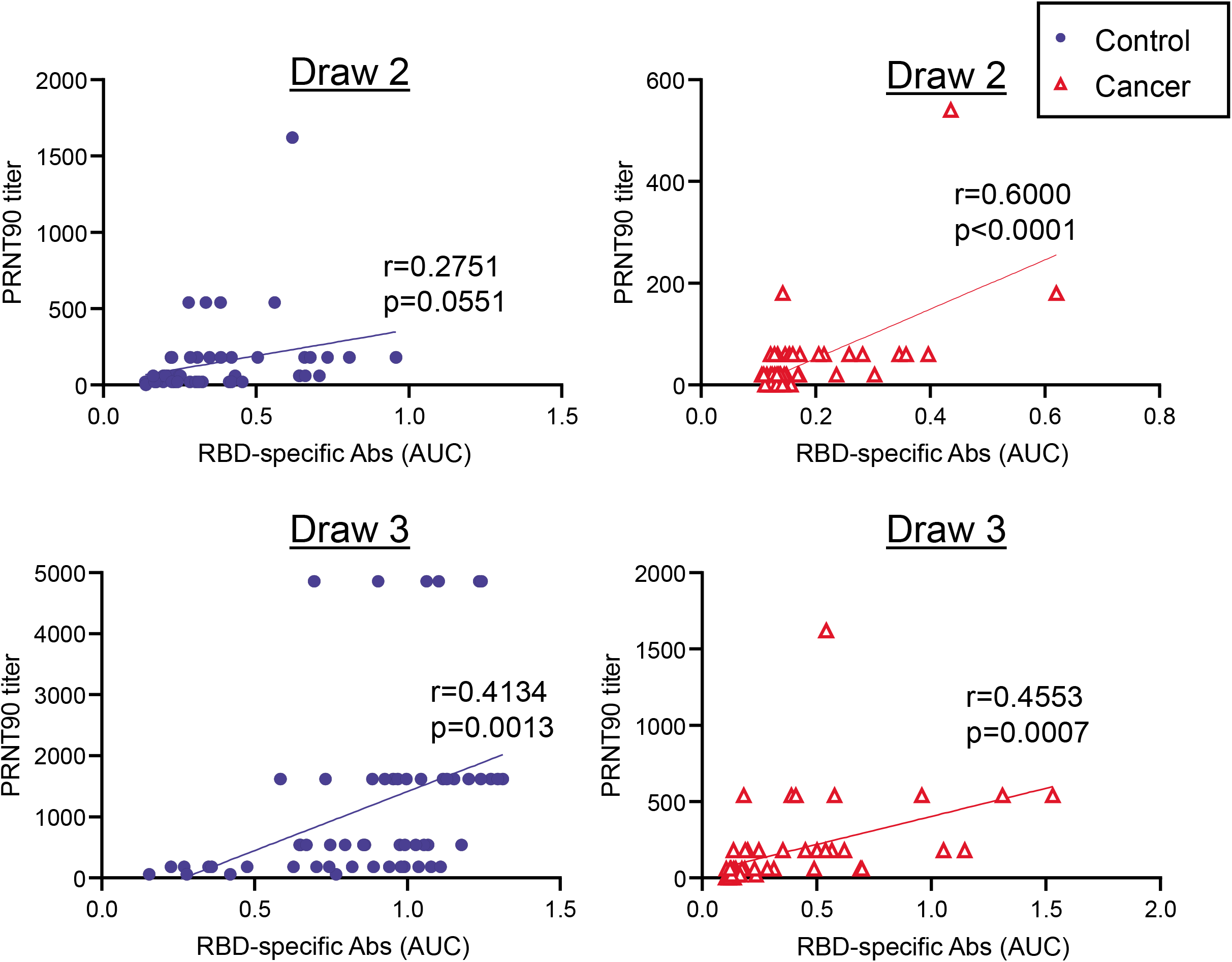
Correlation between RBD-binding antibodies and virus-neutralization. RBD titers were plotted against PRNT levels for control and cancer cohorts at draws 2 and 3. Pearson correlation analyses were performed.

**Extended Data Figure 4:**
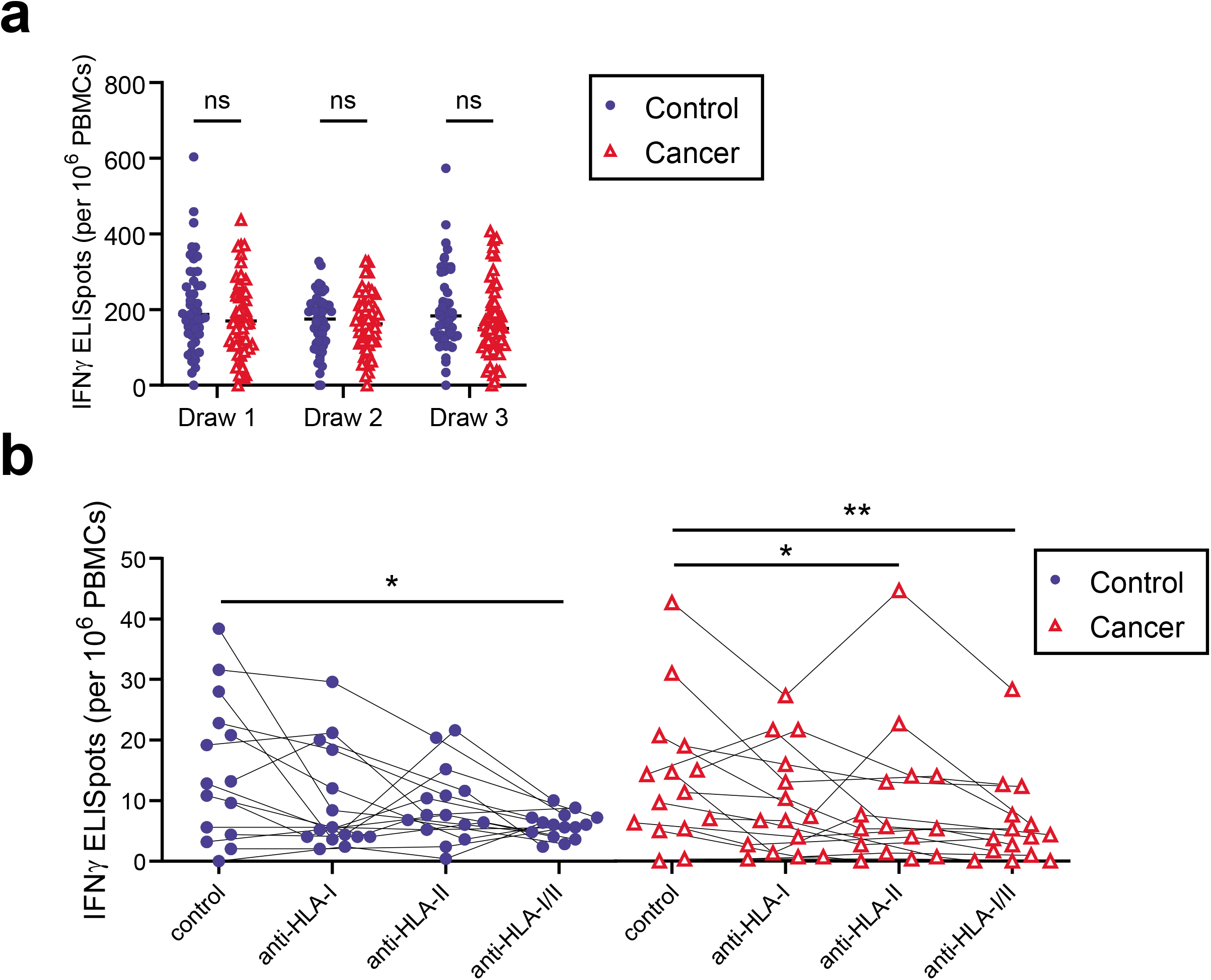
T cell activation in control and cancer cohorts. **a**, PBMCs were cultured for 24 h in the presence of an activating anti-CD3 antibody. IFN*γ*-producing cells were quantified by ELISPOT. P-values were calculated by 2-way ANOVA in which individual samples across blood draws were paired. **b,** Spike-specific T cell activation was quantified in the presence or absence of anti-HLA-I and/or anti-HLA-II blocking antibodies. P-values were calculated using 2-way ANOVA with post-hoc Tukey’s multiple comparisons test.

**Extended Data Figure 5:**
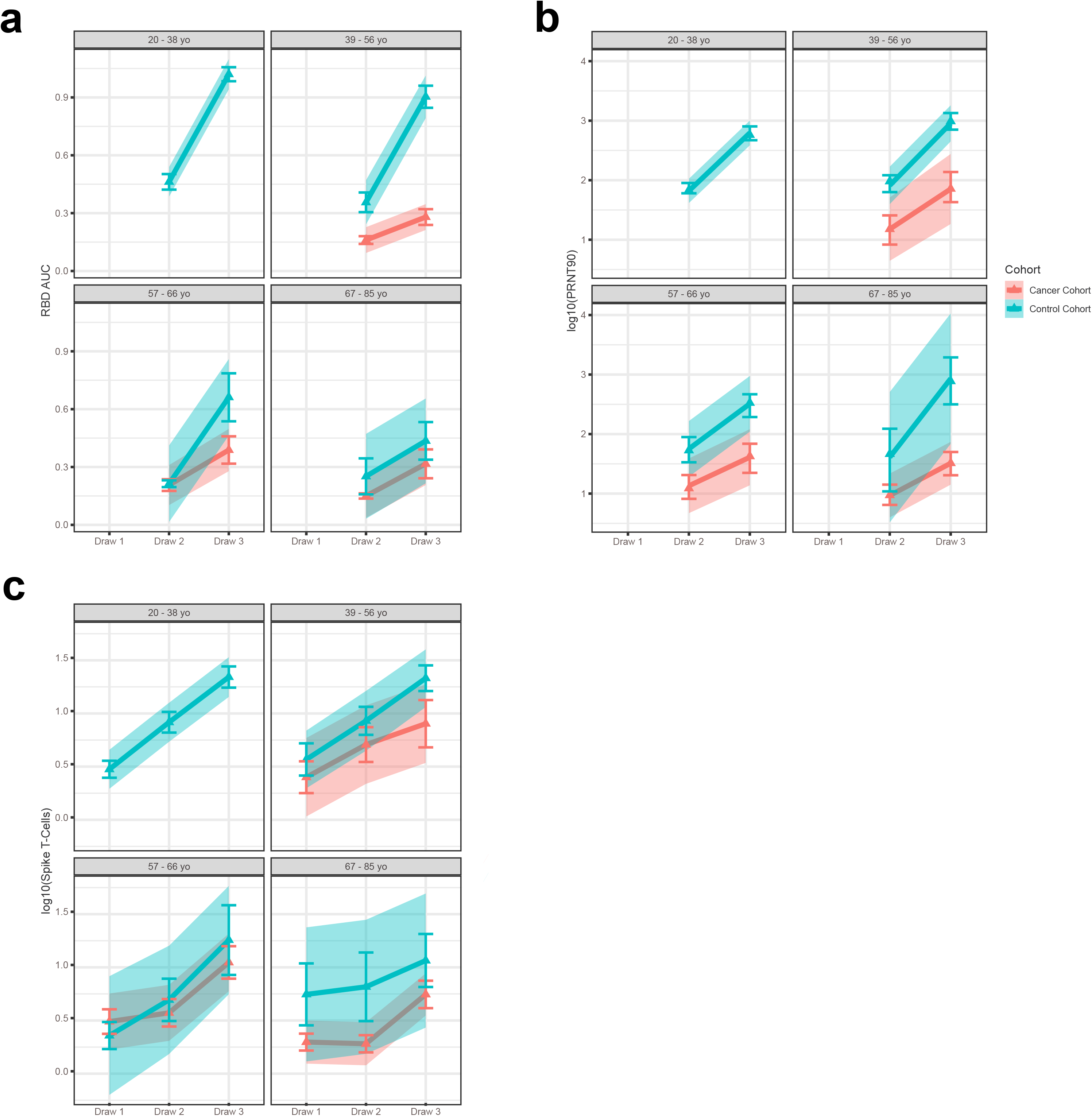
**a,** Trajectory between two draws for RBD AUC for each cohort stratified by age quartile. **b**, Trajectory between two draws for log_10_(PRNT90) for each cohort stratified by age quartile. **c**, Trajectory between three draws for Spike-specific T cell frequencies for each cohort stratified by age quartile. RBD AUC and cohort differences were moderated by age (p-value = 0.01). This effect was driven by the effect of age on the control cohort, increasing age was associated with lower RBD AUC, while the cancer cohort levels were similar across the three upper age quartiles. The difference between the cancer cohort and control cohort was different at draw 3 for all age quartiles 2 - 4. There was no statistically significant difference in the relationship between log_10_(PRNT90) or Spike-specific T-cell frequencies by age. There was a degree of variability in Spike-specific T-cell frequency measurements in both cohorts though the trend in lower draw 3 measures in the cancer cohort remained consistent.

**Extended Data Figure 6:**
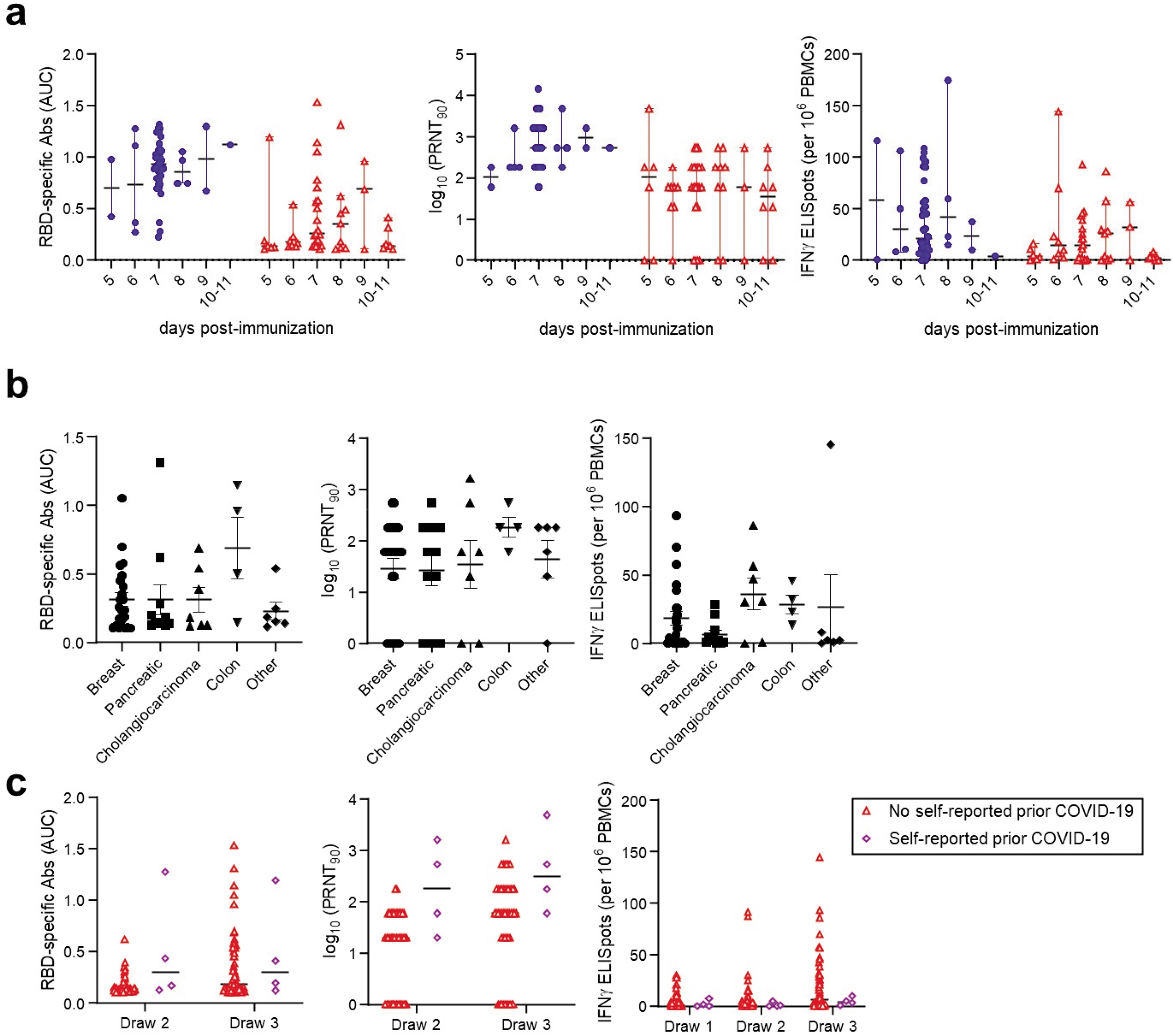
Immune responses grouped by time post-vaccination or tumor type. **a,** RBD-specific antibodies, neutralizing titers, and Spike-specific T cells were plotted as a function of time after the second vaccination. P-values were calculated within each cohort using 1-way ANOVA with post-hoc Sidek’s multiple comparisons test. No significant differences were observed. **b,** RBD-specific antibodies, neutralizing titers, and Spike-specific T cells were plotted as a function of tumor type. P-values were calculated using 1-way ANOVA with post-hoc Sidek’s multiple comparisons test. No significant differences were observed. **c**, RBD-specific antibodies, neutralizing titers, and Spike-specific T cells were plotted comparing participants who either did or did not self-report prior COVID-19.

**Extended Data Figure 7:**
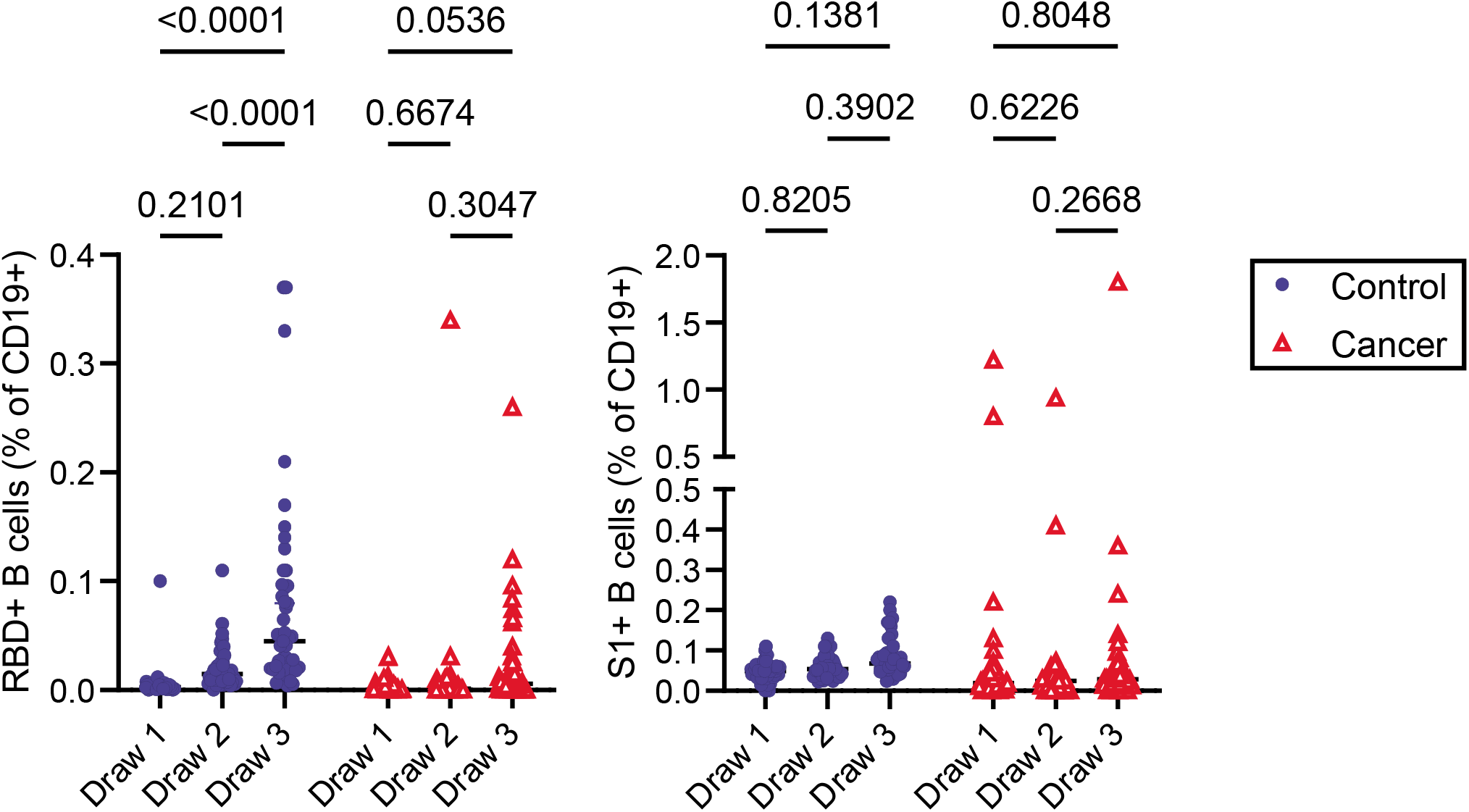
Quantification of RBD- and S1-specific B cells after vaccination. RBD- and S1-specific CD19+ B cell frequencies were measured using gating strategies shown in Extended Data Figure 1 and Figure 4a. Cells that bind both RBD and S1 are annotated as RBD+, whereas cells that are specific only for S1 are denoted as S1+. P-values were calculated using paired 2-way ANOVA.

**Extended Data Figure 8:**
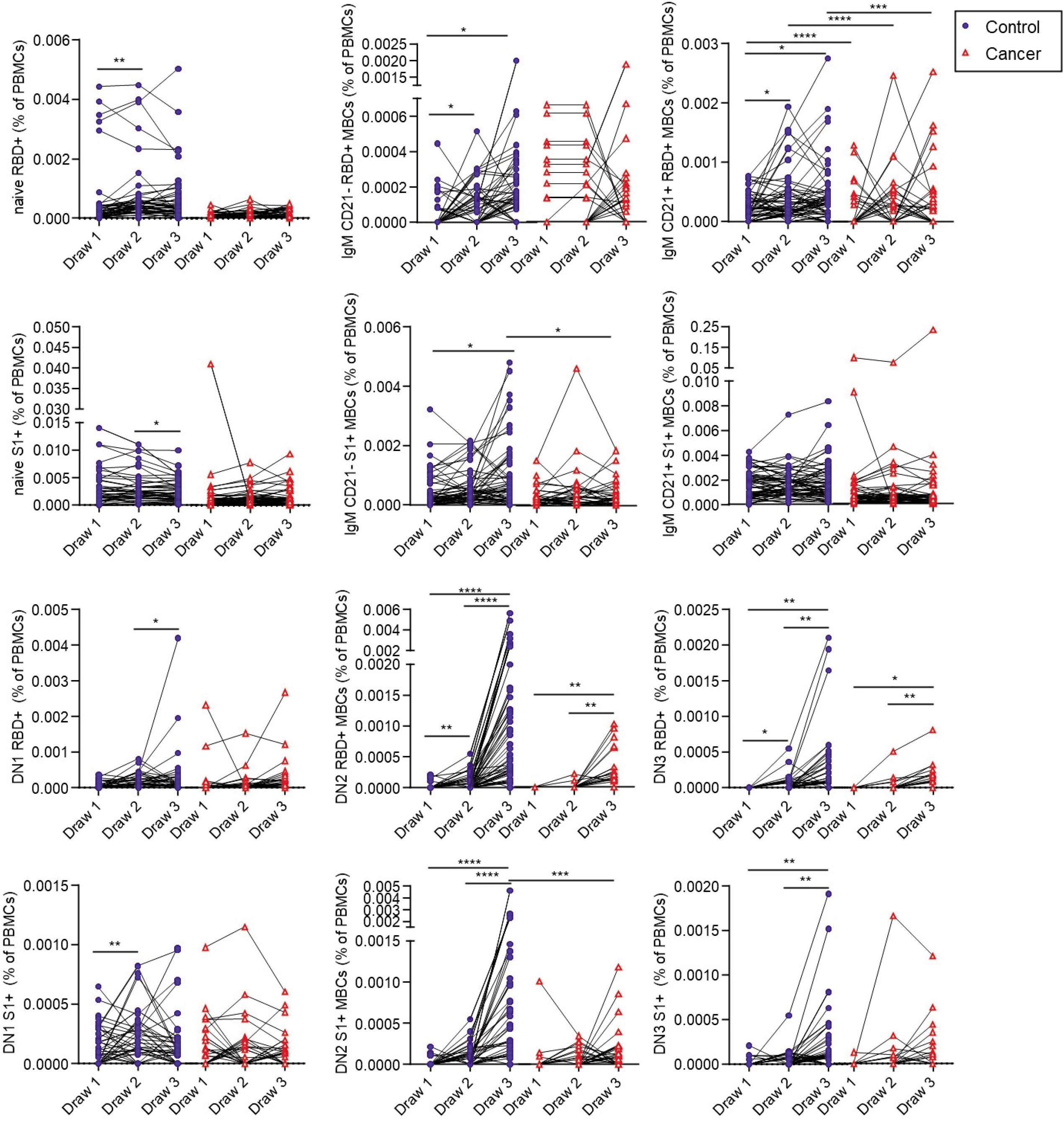
Quantification of memory B cell subsets after vaccination. Cells that bind both RBD and S1 are annotated as RBD+, whereas cells that are specific only for S1 are denoted as S1+. Lines connect the same individual across blood draws, analyses were done on the arcsin of the square-root transformation, to standardize the small percentages. There is a statistically significant difference in slopes between cancer and control cohorts for RBD+ and S1+ (p < 0.0001 and < 0.0001, respectively) and the average rate of change is increasing in the control cohort for both RBD+ and S1+, though the magnitude is only statistically significant between draws 2 and 3 for RBD+ between the cancer cohort and the control cohort (p=0.5806 and p< 0.0001, respectfully) and S1+ (p = 0.3511 and < 0.0001, respectively) P-values were calculated by repeated measures ANOVA. *p<0.05; **p<0.01; ***p<0.001; ****p<0.0001

**Extended Data Figure 9:**
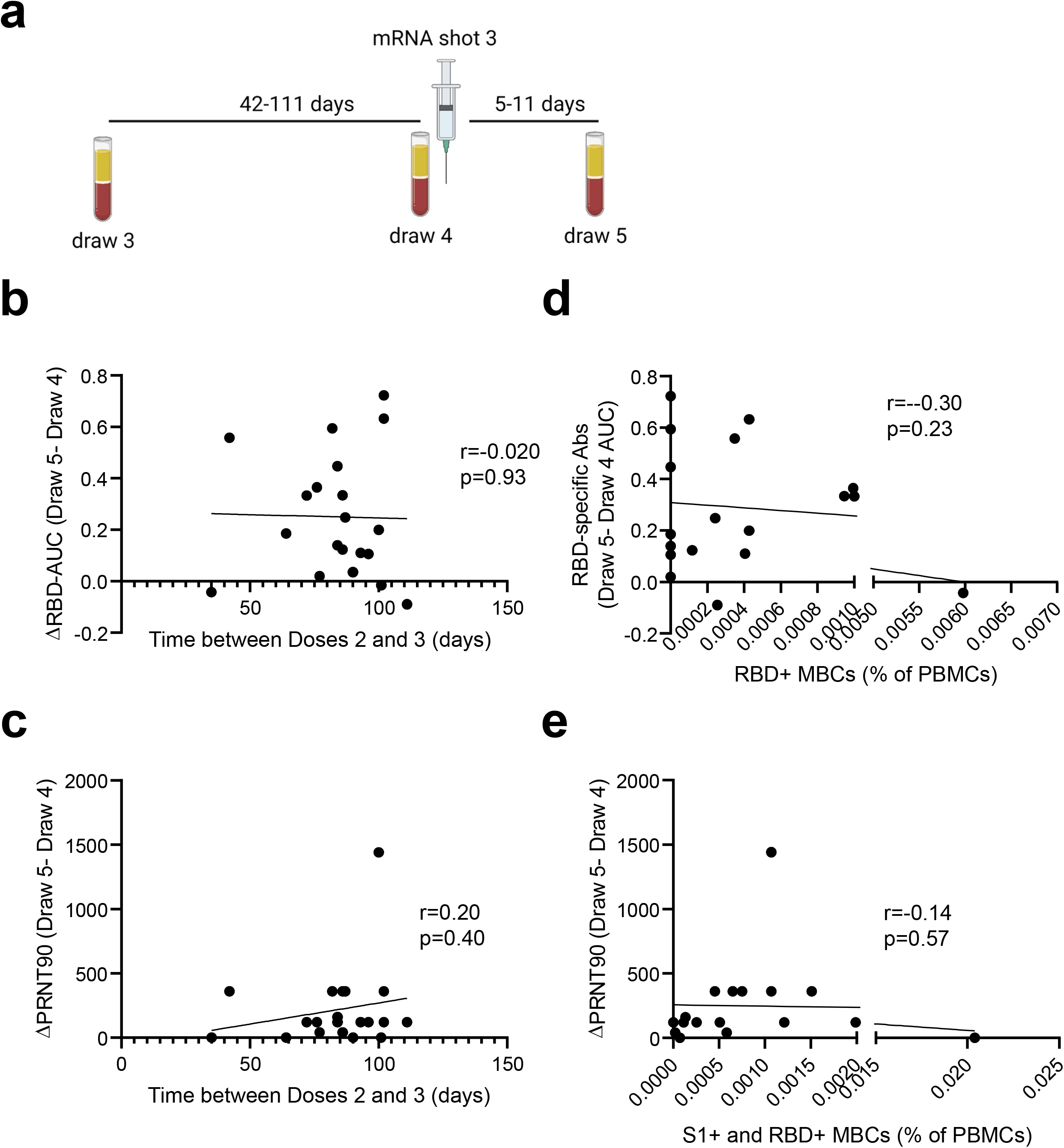
Correlation between memory B cells and anamnestic antibody responses. **a,** RBD-specific memory B cell frequencies at Draw 3 (calculated as in Figure 4) were plotted against the difference in RBD antibodies between Draws 4 and 5. Simple linear regression was performed. **b,** RBD- and S1-specific memory B cell frequencies at Draw 3 (calculated as in Figure 4) were plotted against the difference in PRNT-90 titers between Draws 4 and 5. Simple linear regression was performed.

**Extended Data Figure 10:**
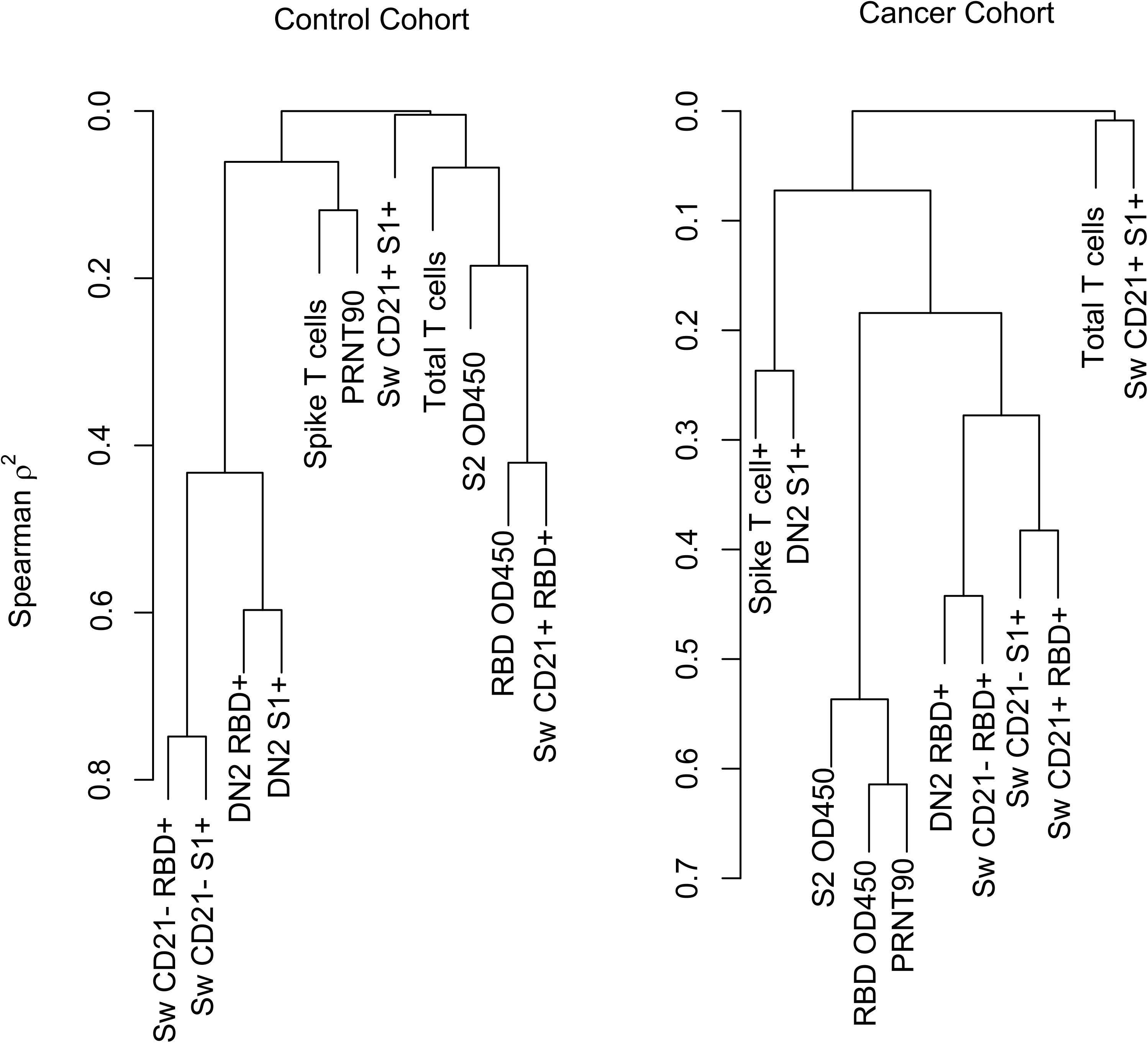
Hierarchical clustering at the variable level, using Spearman’s rank order statistic was performed to evaluate both the correlation (simularity) of immune biomarkers after they are grouped into similar clusters. Of note is the different pattern of both clustering and similarity of the clustered variables between the control and cancer cohorts. Specifically, in the control cohort the b-cell data cluster together into two clusters (including both switched CD21+ and DN2) with a high degree of correlation (spearman correlation of 0.80) a pattern that was not seen in the cancer cohort with the only obvious cluster was the neut titers, RBD and S2 OD -- with a correlation of 0.6.

**Extended Data Figure 11:**
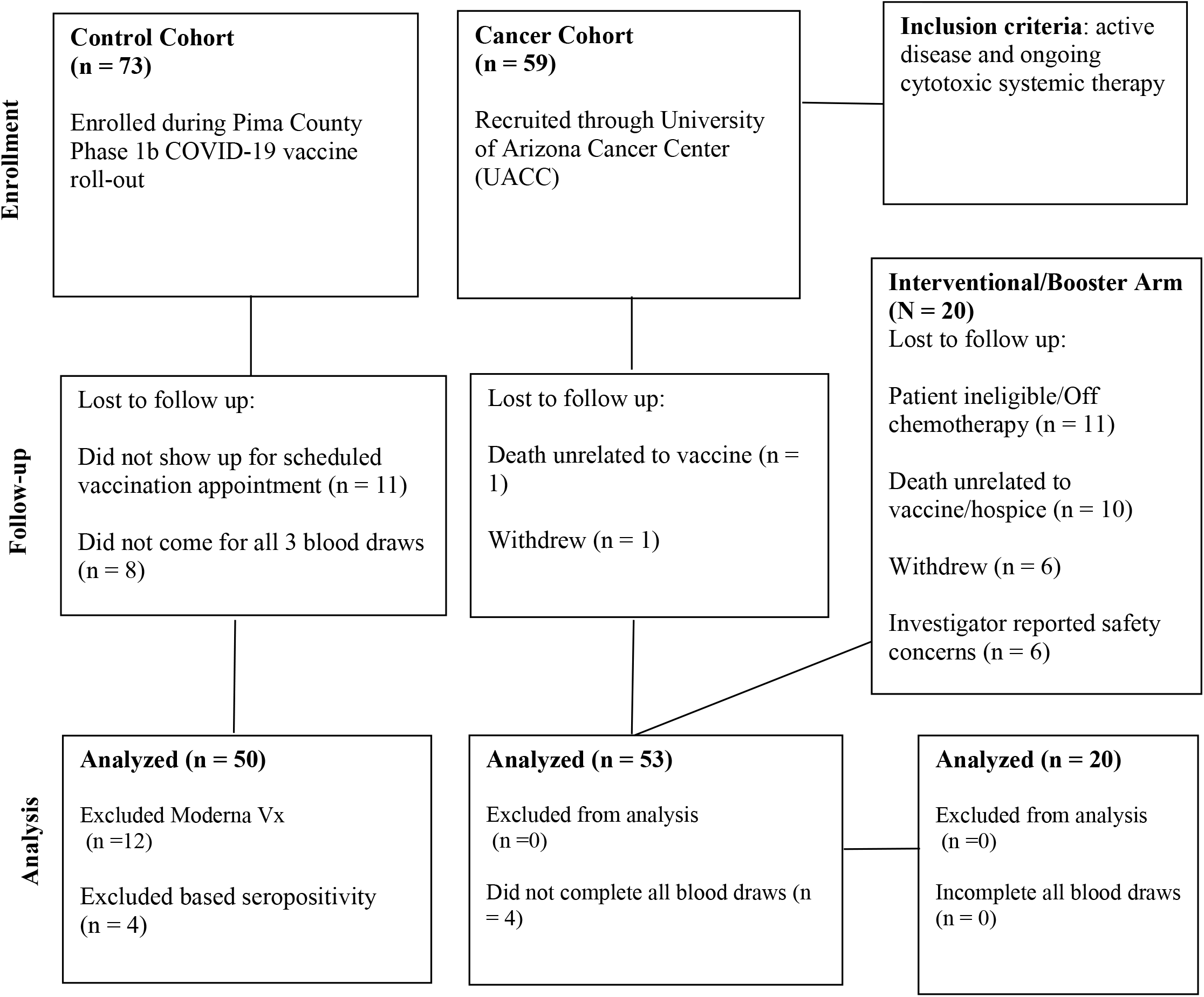
CONSORT flow diagram.

**Extended Data Table 1.** Chemotherapy Regimen and Groupings.

| Chemotherapy Regimen | Cancer Cohort (N) | Chemotherapy Group | Booster Group (N) |
| --- | --- | --- | --- |
| 5-FU, Irinotecan, and Leucovorin | 3 | Fluoropyrimidine-based | 2 |
| 5-FU/Leucovorin | 1 | Fluoropyrimidine-based | 1 |
| Nab-paclitaxel | 2 | Taxane/other antimicrotubule-based |  |
| AIM | 1 | Anthracycline-based |  |
| Avastin and Navelbine | 1 | Taxane/other antimicrotubule-based |  |
| Capecitabine | 3 | Fluoropyrimidine-based |  |
| Capecitabine + Bevacizumab | 1 | Fluoropyrimidine-based | 2 |
| Doxil | 1 | Anthracycline-based |  |
| Enhertu | 1 | Other Targeted Cytotoxics | 1 |
| Eribulin | 1 | Taxane/other antimicrotubule-based |  |
| Faslodex + Abemaciclib | 1 | Oral CDK4/6-based |  |
| Faslodex + Palbociclib | 1 | Oral CDK4/6-based | 1 |
| FLOT | 1 | Fluoropyrimidine-based |  |
| Folfirinox | 2 | Fluoropyrimidine-based | 1 |
| Folfirinox + CPI-613 | 1 | Fluoropyrimidine-based | 1 |
| FOLFOX | 1 | Fluoropyrimidine-based |  |
| FOLFOX + Bevacizumab | 1 | Fluoropyrimidine-based |  |
| Fulvestrant + Palbociclib | 1 | Oral CDK4/6-based |  |
| Gem/cis/nab-paclitaxel | 5 | Gemcitabine-based | 5 |
| Gemcitabine and nab-paclitaxel | 4 | Gemcitabine-based | 3 |
| Gemcitabine and Cisplatin | 4 | Gemcitabine-based | 2 |
| Ibrance and Anastrozole | 1 | Oral CDK4/6-based |  |
| Letrozole + Ibrance | 2 | Oral CDK4/6-based |  |
| Paclitaxel and Carboplatin | 2 | Taxane/other antimicrotubule-based |  |
| Palbociclib | 1 | Oral CDK4/6-based | 2 |
| Sacituzumab govitecan | 1 | Other Targeted Cytotoxics |  |
| Stivarga | 1 | Other Targeted Cytotoxics | 1 |
| T-DM1 + Palbo | 1 | Oral CDK4/6-based |  |
| Taxol | 3 | Taxane/other antimicrotubule-based |  |
| Topotecan + Bevacizumab | 1 | Other Targeted Cytotoxics |  |
| ZN-C5 and Palbociclib | 2 | Oral CDK4/6-based |  |

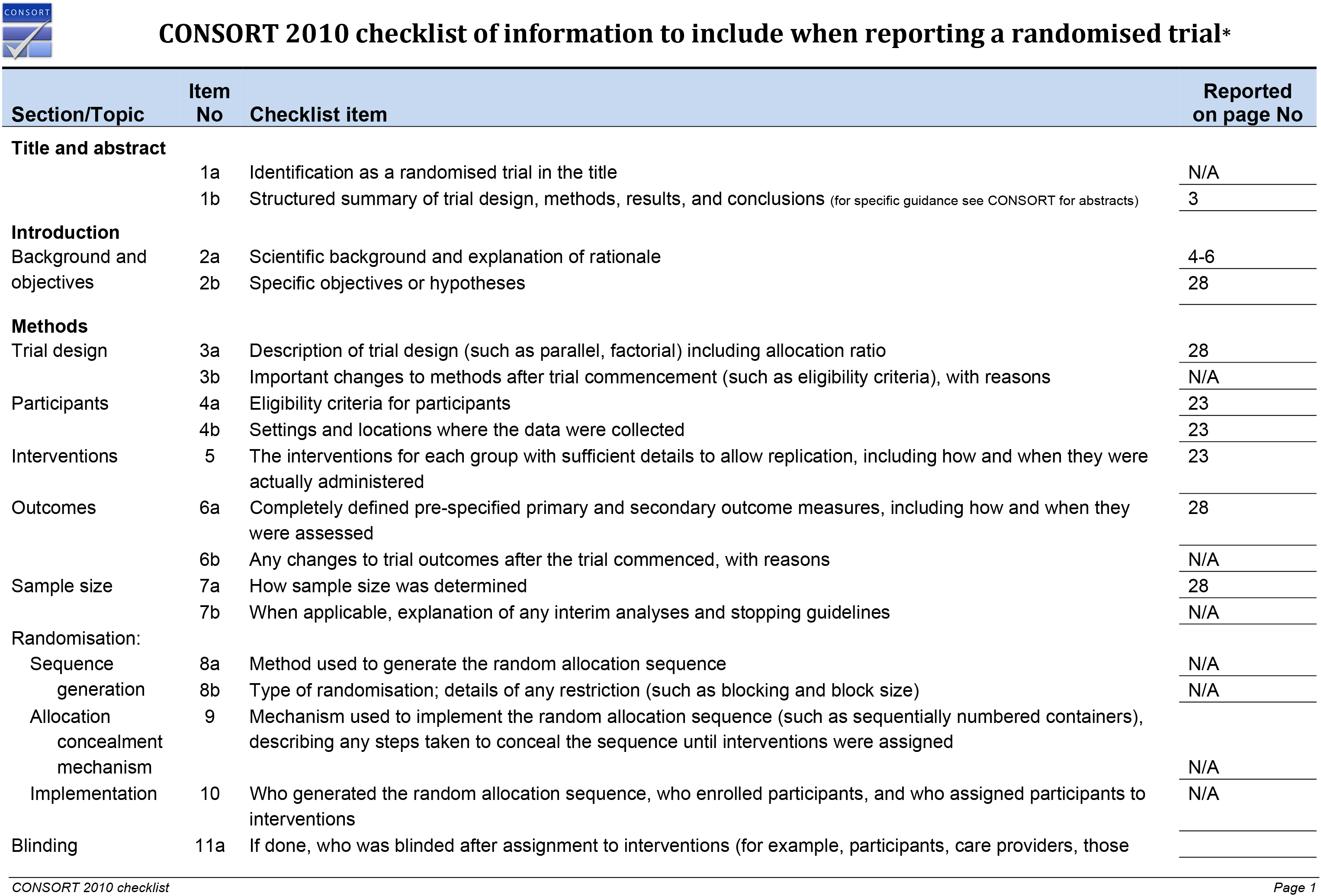
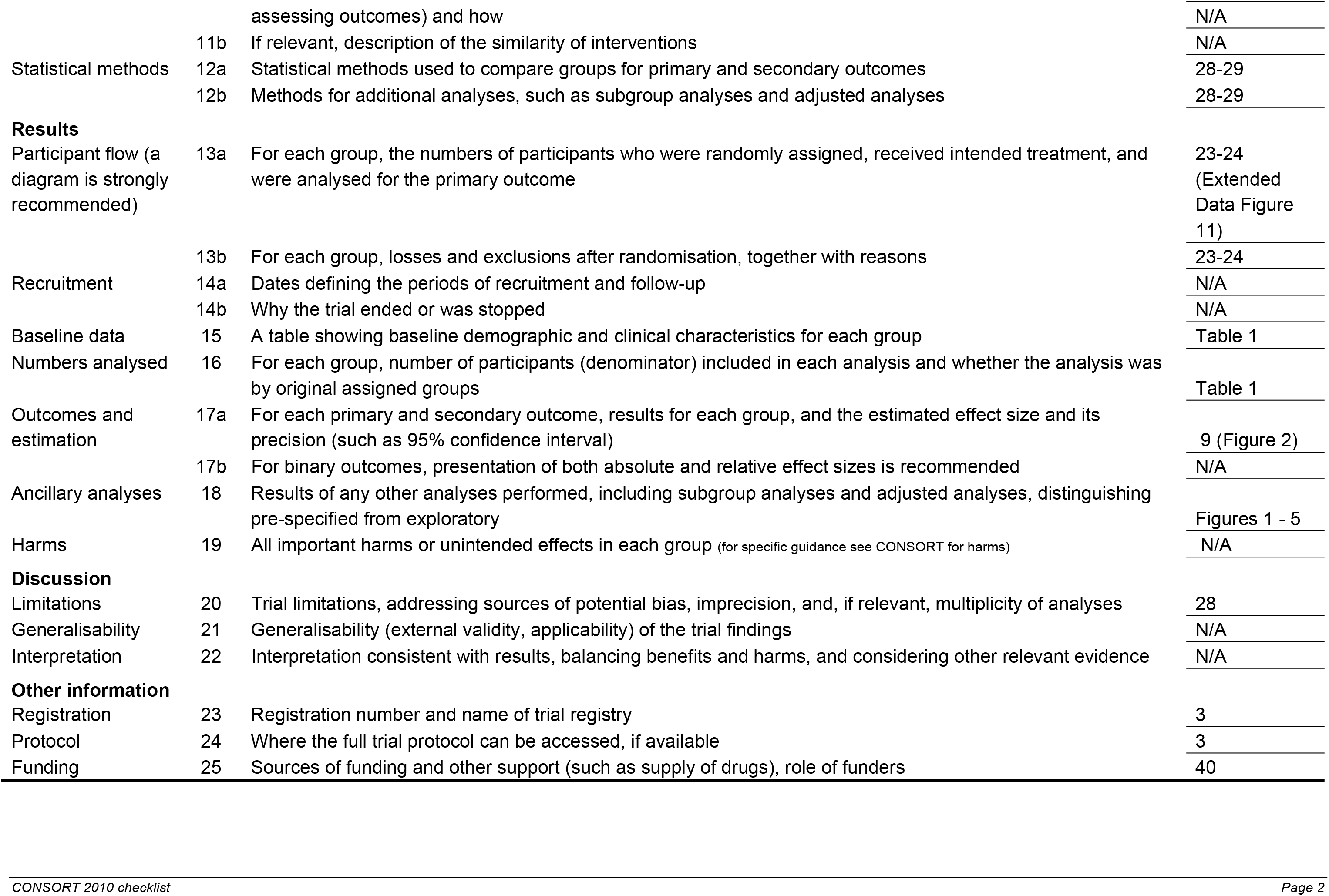
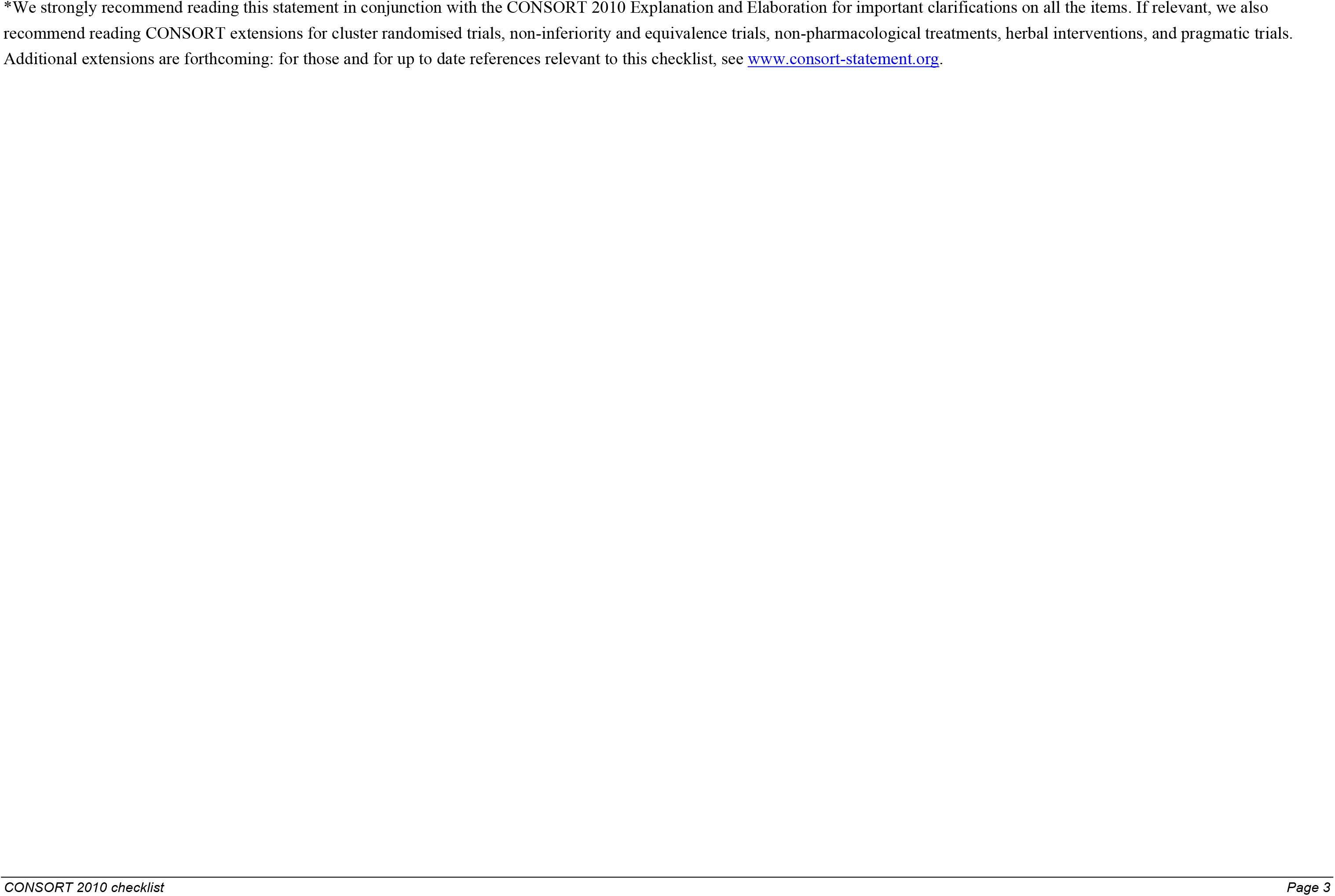

## References

1. Wu, F. et al. A new coronavirus associated with human respiratory disease in China. Nature 579, 265–269 (2020).

2. Zhou, P. et al. A pneumonia outbreak associated with a new coronavirus of probable bat origin. Nature 579, 270–273 (2020).

3. Polack, F. P. et al. Safety and Efficacy of the BNT162b2 mRNA Covid-19 Vaccine. N. Engl. J. Med. 0, null (2020).

4. Baden, L. R. et al. Efficacy and Safety of the mRNA-1273 SARS-CoV-2 Vaccine. N. Engl. J. Med. 384, 403–416 (2021).

5. Thompson, M. G. et al. Interim Estimates of Vaccine Effectiveness of BNT162b2 and mRNA-1273 COVID-19 Vaccines in Preventing SARS-CoV-2 Infection Among Health Care Personnel, First Responders, and Other Essential and Frontline Workers - Eight U.S. Locations, December 2020-March 2021. MMWR Morb. Mortal. Wkly. Rep. 70, 495–500 (2021).

6. Abu-Raddad, L. J., Chemaitelly, H. & Butt, A. A. Effectiveness of the BNT162b2 Covid-19 Vaccine against the B.1.1.7 and B.1.351 Variants. N. Engl. J. Med. 0, null (2021).

7. Dagan, N. et al. BNT162b2 mRNA Covid-19 Vaccine in a Nationwide Mass Vaccination Setting. N. Engl. J. Med. 384, 1412–1423 (2021).

8. Levine-Tiefenbrun, M. et al. Initial report of decreased SARS-CoV-2 viral load after inoculation with the BNT162b2 vaccine. Nat. Med. 1–3 (2021) doi:10.1038/s41591-021-01316-7.

9. . Kuderer, N. M. et al. Clinical impact of COVID-19 on patients with cancer (CCC19): a cohort study. Lancet Lond. Engl. 395, 1907–1918 (2020).

10. Kemp, S. A. et al. SARS-CoV-2 evolution during treatment of chronic infection. Nature 592, 277–282 (2021).

11. McCarthy, K. R. et al. Recurrent deletions in the SARS-CoV-2 spike glycoprotein drive antibody escape. Science 371, 1139–1142 (2021).

12. Avanzato, V. A. et al. Case Study: Prolonged Infectious SARS-CoV-2 Shedding from an Asymptomatic Immunocompromised Individual with Cancer. Cell 183, 1901–1912.e9 (2020).

13. Aydillo, T. et al. Shedding of Viable SARS-CoV-2 after Immunosuppressive Therapy for Cancer. N. Engl. J. Med. 383, 2586–2588 (2020).

14. Choi, B. et al. Persistence and Evolution of SARS-CoV-2 in an Immunocompromised Host. N. Engl. J. Med. 383, 2291–2293 (2020).

15. Hensley, M. K. et al. Intractable COVID-19 and Prolonged SARS-CoV-2 Replication in a CAR-T-cell Therapy Recipient: A Case Study. Clin. Infect. Dis. Off. Publ. Infect. Dis. Soc. Am. (2021) doi:10.1093/cid/ciab072.

16. Cobey, S., Larremore, D. B., Grad, Y. H. & Lipsitch, M. Concerns about SARS-CoV-2 evolution should not hold back efforts to expand vaccination. Nat. Rev. Immunol. 21, 330–335 (2021).

17. Khoury, D. S. et al. What level of neutralising antibody protects from COVID-19? medRxiv 2021.03.09.21252641 (2021) doi:10.1101/2021.03.09.21252641.

18. Herishanu, Y. et al. Efficacy of the BNT162b2 mRNA COVID-19 vaccine in patients with chronic lymphocytic leukemia. Blood 137, 3165–3173 (2021).

19. Deepak, P. et al. Glucocorticoids and B Cell Depleting Agents Substantially Impair Immunogenicity of mRNA Vaccines to SARS-CoV-2. medRxiv 2021.04.05.21254656 (2021) doi:10.1101/2021.04.05.21254656.

20. Boyarsky, B. J. et al. Immunogenicity of a Single Dose of SARS-CoV-2 Messenger RNA Vaccine in Solid Organ Transplant Recipients. JAMA 325, 1784–1786 (2021).

21. Boyarsky, B. J. et al. Antibody Response to 2-Dose SARS-CoV-2 mRNA Vaccine Series in Solid Organ Transplant Recipients. JAMA (2021) doi:10.1001/jama.2021.7489.

22. Monin, L. et al. Safety and immunogenicity of one versus two doses of the COVID-19 vaccine BNT162b2 for patients with cancer: interim analysis of a prospective observational study. Lancet Oncol. S1470204521002138 (2021) doi:10.1016/S1470-2045(21)00213-8.

23. Center for Devices and Radiological Health. EUA Authorized Serology Test Performance. FDA (2020).

24. Goel, R. R. et al. Distinct antibody and memory B cell responses in SARS-CoV-2 naïve and recovered individuals following mRNA vaccination. Sci. Immunol. 6, (2021).

25. Ladner, J. T. et al. Epitope-resolved profiling of the SARS-CoV-2 antibody response identifies cross-reactivity with endemic human coronaviruses. Cell Rep. Med. 2, 100189 (2021).

26. Anderson, E. M. et al. Seasonal human coronavirus antibodies are boosted upon SARS-CoV-2 infection but not associated with protection. Cell 184, 1858–1864.e10 (2021).

27. Nguyen-Contant, P. et al. S Protein-Reactive IgG and Memory B Cell Production after Human SARS-CoV-2 Infection Includes Broad Reactivity to the S2 Subunit. mBio 11, (2020).

28. Shrock, E. et al. Viral epitope profiling of COVID-19 patients reveals cross-reactivity and correlates of severity. Science 370, (2020).

29. Ng, K. W. et al. Preexisting and de novo humoral immunity to SARS-CoV-2 in humans. Science 370, 1339–1343 (2020).

30. Song, G. et al. Cross-reactive serum and memory B cell responses to spike protein in SARS-CoV-2 and endemic coronavirus infection. bioRxiv 2020.09.22.308965 (2020) doi:10.1101/2020.09.22.308965.

31. Greaney, A. J. et al. The SARS-CoV-2 mRNA-1273 vaccine elicits more RBD-focused neutralization, but with broader antibody binding within the RBD. bioRxiv 2021.04.14.439844 (2021) doi:10.1101/2021.04.14.439844.

32. Piccoli, L. et al. Mapping Neutralizing and Immunodominant Sites on the SARS-CoV-2 Spike Receptor-Binding Domain by Structure-Guided High-Resolution Serology. Cell 183, 1024–1042.e21 (2020).

33. Zinkernagel, R. M. & Hengartner, H. Protective ‘immunity’ by pre-existent neutralizing antibody titers and preactivated T cells but not by so-called ‘immunological memory’. Immunol. Rev. 211, 310–319 (2006).

34. Abayasingam, A. et al. Long-term persistence of RBD+ memory B cells encoding neutralizing antibodies in SARS-CoV-2 infection. Cell Rep. Med. 2, 100228 (2021).

35. Ripperger, T. J. et al. Orthogonal SARS-CoV-2 Serological Assays Enable Surveillance of Low-Prevalence Communities and Reveal Durable Humoral Immunity. Immunity 53, 925–933.e4 (2020).

36. Wec, A. Z. et al. Broad neutralization of SARS-related viruses by human monoclonal antibodies. Science 369, 731–736 (2020).

37. Chi, X. et al. A neutralizing human antibody binds to the N-terminal domain of the Spike protein of SARS-CoV-2. Science (2020) doi:10.1126/science.abc6952.

38. Sekine, T. et al. Robust T Cell Immunity in Convalescent Individuals with Asymptomatic or Mild COVID-19. Cell 183, 158–168.e14 (2020).

39. Rydyznski Moderbacher, C. et al. Antigen-Specific Adaptive Immunity to SARS-CoV-2 in Acute COVID-19 and Associations with Age and Disease Severity. Cell 183, 996–1012.e19 (2020).

40. Ogbe, A. et al. T cell assays differentiate clinical and subclinical SARS-CoV-2 infections from cross-reactive antiviral responses. Nat. Commun. 12, 2055 (2021).

41. McMahan, K. et al. Correlates of protection against SARS-CoV-2 in rhesus macaques. Nature 590, 630–634 (2021).

42. Reynolds, C. J. et al. Discordant neutralizing antibody and T cell responses in asymptomatic and mild SARS-CoV-2 infection. Sci. Immunol. 5, (2020).

43. Anderson, E. J. et al. Safety and Immunogenicity of SARS-CoV-2 mRNA-1273 Vaccine in Older Adults. N. Engl. J. Med. 383, 2427–2438 (2020).

44. Rodda, L. B. et al. Functional SARS-CoV-2-Specific Immune Memory Persists after Mild COVID-19. Cell 184, 169–183.e17 (2021).

45. Sahin, U. et al. COVID-19 vaccine BNT162b1 elicits human antibody and T H 1 T cell responses. Nature 586, 594–599 (2020).

46. Agerer, B. et al. SARS-CoV-2 mutations in MHC-I-restricted epitopes evade CD8+ T cell responses. Sci. Immunol. 6, (2021).

47. Dan, J. M. et al. Immunological memory to SARS-CoV-2 assessed for up to 8 months after infection. Science (2021) doi:10.1126/science.abf4063.

48. McCallum, M. et al. N-terminal domain antigenic mapping reveals a site of vulnerability for SARS-CoV-2. Cell 184, 2332–2347.e16 (2021).

49. Dogan, I. et al. Multiple layers of B cell memory with different effector functions. Nat. Immunol. 10, 1292–1299 (2009).

50. Pape, K. A., Taylor, J. J., Maul, R. W., Gearhart, P. J. & Jenkins, M. K. Different B cell populations mediate early and late memory during an endogenous immune response. Science 331, 1203–7 (2011).

51. Zuccarino-Catania, G. V. et al. CD80 and PD-L2 define functionally distinct memory B cell subsets that are independent of antibody isotype. Nat. Immunol. 15, 631–637 (2014).

52. Seifert, M. et al. Functional capacities of human IgM memory B cells in early inflammatory responses and secondary germinal center reactions. Proc. Natl. Acad. Sci. 112, E546–E555 (2015).

53. Turner, J. S. et al. Human germinal centres engage memory and naive B cells after influenza vaccination. Nature 586, 127–132 (2020).

54. Lau, D. et al. Low CD21 expression defines a population of recent germinal center graduates primed for plasma cell differentiation. Sci. Immunol. 2, eaai8153 (2017).

55. Jenks, S. A. et al. Distinct Effector B Cells Induced by Unregulated Toll-like Receptor 7 Contribute to Pathogenic Responses in Systemic Lupus Erythematosus. Immunity 49, 725–739.e6 (2018).

56. Knox, J. J. et al. T-bet+ B cells are induced by human viral infections and dominate the HIV gp140 response. JCI Insight 2,.

57. Kyu, S. Y. et al. Frequencies of human influenza-specific antibody secreting cells or plasmablasts post vaccination from fresh and frozen peripheral blood mononuclear cells. J. Immunol. Methods 340, 42–47 (2009).

58. Amanna, I. J. Duration of Humoral Immunity to Common Viral and Vaccine Antigens. N. Engl. J. Med. 357, 1903–1915 (2007).

59. Lavinder, J. J. et al. Identification and characterization of the constituent human serum antibodies elicited by vaccination. Proc. Natl. Acad. Sci. U. S. A. (2014) doi:10.1073/pnas.1317793111.

60. Purtha, W. E., Tedder, T. F., Johnson, S., Bhattacharya, D. & Diamond, M. S. Memory B cells, but not long-lived plasma cells, possess antigen specificities for viral escape mutants. J. Exp. Med. 208, 2599–2606 (2011).

61. Wong, R. et al. Affinity-Restricted Memory B Cells Dominate Recall Responses to Heterologous Flaviviruses. Immunity 53, 1078–1094.e7 (2020).

62. Smith, K. G. C., Light, A., Nossal, G. J. V. & Tarlinton, D. M. The extent of affinity maturation differs between the memory and antibody-forming cell compartments in the primary immune response. EMBO J. 16, 2996–3006 (1997).

63. Angyal, A. et al. T-Cell and Antibody Responses to First BNT162b2 Vaccine Dose in Previously SARS-CoV-2-Infected and Infection-Naive UK Healthcare Workers: A Multicentre, Prospective, Observational Cohort Study. https://papers.ssrn.com/abstract=3820576 (2021) doi:10.2139/ssrn.3820576.

64. Wumkes, M. L. et al. Serum antibody response to influenza virus vaccination during chemotherapy treatment in adult patients with solid tumours. Vaccine 31, 6177–6184 (2013).

65. Puthillath, A. et al. Serological immune responses to influenza vaccine in patients with colorectal cancer. Cancer Chemother. Pharmacol. 67, 111–115 (2011).

66. Robbiani, D. F. et al. Convergent antibody responses to SARS-CoV-2 in convalescent individuals. Nature 584, 437–442 (2020).

67. Lumley, S. F. et al. Antibody Status and Incidence of SARS-CoV-2 Infection in Health Care Workers. N. Engl. J. Med. 384, 533–540 (2021).

68. Abu-Raddad, L. J. et al. Assessment of the risk of SARS-CoV-2 reinfection in an intense re-exposure setting. Clin. Infect. Dis. Off. Publ. Infect. Dis. Soc. Am. (2020) doi:10.1093/cid/ciaa1846.

69. Abu-Raddad, L. J. et al. SARS-CoV-2 antibody-positivity protects against reinfection for at least seven months with 95% efficacy. EClinicalMedicine 35, (2021).

70. Long, Q.-X. et al. Clinical and immunological assessment of asymptomatic SARS-CoV-2 infections. Nat. Med. 10.1038/s41591-020-0965-6, (2020).

71. Liao, M. et al. Single-cell landscape of bronchoalveolar immune cells in patients with COVID-19. Nat. Med. 26, 842–844 (2020).

72. Mateus, J. et al. Selective and cross-reactive SARS-CoV-2 T cell epitopes in unexposed humans. Science 370, 89–94 (2020).

73. Swadling, L. et al. Pre-existing polymerase-specific T cells expand in abortive seronegative SARS-CoV-2 infection. medRxiv 2021.06.26.21259239 (2021) doi:10.1101/2021.06.26.21259239.

74. Zhou, D. et al. Evidence of escape of SARS-CoV-2 variant B.1.351 from natural and vaccine-induced sera. Cell 184, 2348–2361.e6 (2021).

75. Edara, V.-V. et al. Infection and Vaccine-Induced Neutralizing-Antibody Responses to the SARS-CoV-2 B.1.617 Variants. N. Engl. J. Med. 0, null (2021).

76. Werbel, W. A. et al. Safety and Immunogenicity of a Third Dose of SARS-CoV-2 Vaccine in Solid Organ Transplant Recipients: A Case Series. Ann. Intern. Med. (2021) doi:10.7326/L21-0282.

77. Kamar, N. et al. Three Doses of an mRNA Covid-19 Vaccine in Solid-Organ Transplant Recipients. N. Engl. J. Med. 0, null (2021).

78. Khoury, D. S. et al. Neutralizing antibody levels are highly predictive of immune protection from symptomatic SARS-CoV-2 infection. Nat. Med. 1–7 (2021) doi:10.1038/s41591-021-01377-8.

79. Krammer, F. et al. Antibody Responses in Seropositive Persons after a Single Dose of SARS-CoV-2 mRNA Vaccine. N. Engl. J. Med. 384, 1372–1374 (2021).

80. Andrews, S. F. et al. High Preexisting Serological Antibody Levels Correlate with Diversification of the Influenza Vaccine Response. J. Virol. 89, 3308–3317 (2015).

